# Independent and combined effects of nutrition and sanitation interventions on enteric pathogen carriage and child growth in rural Cambodia: a factorial cluster-randomised controlled trial

**DOI:** 10.1101/2021.05.21.21257546

**Authors:** Amanda Lai, Irene Velez, Ramya Ambikapathi, Krisna Seng, Karen Levy, Erin Kowalsky, David Holcomb, Konstantinos T. Konstantinidis, Oliver Cumming, Joe Brown

## Abstract

**Background:** Childhood exposure to enteric pathogens associated with poor sanitation contributes to undernutrition, associated with adverse effects later in life. This trial assessed the independent and combined effects of nutrition and sanitation interventions on child growth outcomes and enteric pathogen infection in rural Cambodia, where the prevalence of childhood stunting remains high.

**Methods:** We conducted a factorial cluster-randomised controlled trial of 4,015 households with 4,124 children (1-28 months of age at endline) across three rural provinces in Cambodia. Fifty-five communes (clusters) were randomly assigned to a control arm or one of three treatments: a nutrition-only arm, a sanitation-only arm, and a combined nutrition and sanitation arm receiving both treatments. The primary outcome was length-for-age Z-score (LAZ); other outcomes included weight-for-age Z-score (WAZ), weight-for-length Z-score (WLZ), stunting, wasting, underweight, and caregiver-reported diarrhoea. We assayed stool specimens from a subset of all children (n = 1,620) for 27 enteric pathogens (14 bacteria, 6 viruses, 3 protozoa, and 4 soil-transmitted helminths) and estimated effects of interventions on enteric pathogen detection and density. Analysis was by intention-to-treat. The trial was pre-registered with ISRCTN Registry (<u>ISRCTN77820875</u>).

**Findings:** Self-reported adherence was high for the nutrition intervention but uptake was low for sanitation. Compared with a mean LAZ of -1.04 (SD 1.2) in the control arm, children in the nutrition-only arm (LAZ +0.08, 95% CI -0.01-0.18) and combined nutrition and sanitation arm (LAZ +0.10, 95% CI 0.01-0.20) experienced greater linear growth; there were no measurable differences in LAZ in the sanitation-only arm (LAZ -0.05, 95% CI -0.16-0.05). We found no effect of any intervention (delivered independently or combined) on either enteric pathogen frequency or pathogen load in stool. Compared with a mean WAZ of -1.05 (SD 1.1) in the control arm, children in the nutrition-only arm (WAZ +0.10, 95% CI 0.00-0.19) and combined intervention arm (WAZ +0.11, 95% CI 0.03-0.20) were heavier for their age; there was no difference in WAZ in the sanitation-only arm. There were no differences between arms in prevalence of stunting, wasting, underweight status, one-week period prevalence of diarrhoea, pathogen prevalence, or pathogen density in stool.

**Interpretation:** Improvements in child growth in nutrition and combined nutrition and sanitation arms are consistent with previous efficacy trials of combined nutrition and sanitation interventions. We found no evidence that the sanitation intervention alone improved child growth or reduced enteric pathogen detection, having achieved only modest changes in access and use.

**Funding:** United States Agency for International Development (USAID), contracts AID-OAA-M-13-00017 and AID-OAA-TO-16-00016. The contents of this publication are the sole responsibility of the authors and do not necessarily reflect the views of USAID or the United States Government.

## Introduction

Childhood undernutrition is associated with higher susceptibility to infectious disease, reduced cognitive function, and various adverse outcomes later in life^1^. Growth faltering is an effect of chronic undernutrition and tends to manifest in a child’s first two years^2^. Many studies have focused on improving infant and child nutrition to achieve better growth outcomes^3,4^. However, nutrition interventions alone have not been successful in eliminating stunting, suggesting that broader interventions addressing other important factors are needed alongside exclusive breastfeeding and improved nutritional intake^5^.

Reducing early childhood exposure to enteric pathogens through safe water, sanitation, and hygiene (WASH) may complement other interventions by reducing diarrhoeal diseases and environmental enteric dysfunction (EED)^6^—both of which can impact early childhood growth and development^7^. Observational studies have found strong associations between child growth faltering and poor access to sanitation^8^. However, recent randomised controlled trials (RCTs) in Zimbabwe^9^, Bangladesh^3^, and Kenya^4^ that delivered standalone household-level sanitation interventions (not coupled with other nutrition or hygiene interventions) were not found to improve child growth.

The community-led total sanitation (CLTS) framework is an approach to ending open defecation (OD) through behavioural change and collective action rather than through the provision of hardware and materials. CLTS and other rural promotion-based interventions shift the focus from individual and household sanitation practices to a community-level concern over OD by triggering collective behaviour change through powerful emotional drivers such as shame and disgust, as well as positive motivators such as improved health, dignity, and pride. Observational studies in Cambodia^10^ and Ecuador^11^ found higher community-level sanitation coverage to be associated with reduced prevalence of stunting. Despite this, recent RCTs employing promotion-based interventions have found mixed effects on child growth. One trial was found to be successful in improving child growth in Mali^12^, but this effect was not observed in other trials elsewhere^13,14^.

This study contributes to a growing body of literature on the impact of combined nutrition and sanitation interventions on early child growth, caregiver-reported diarrhoea, and detection and quantification of enteric pathogens in stool as a proxy for enteric infection. While diarrhoea has been widely used as a primary outcome measure in WASH studies^3,4,12,15^, recent studies have used stool-based detection of enteric pathogens^15^ and anthropometry measurements^3,4,9^ as primary outcomes that are more objectively measurable and may also broadly indicate health status by capturing cumulative effects of exposures via EED^6^. We used a factorial cluster-randomised controlled trial (cRCT) to assess the independent and combined effects of nutrition and sanitation interventions delivered in the context of a large-scale, USAID-funded rural nutrition and sanitation/hygiene program in Cambodia. We hypothesised that children receiving both sanitation and nutrition interventions would have increased linear growth compared with children from control areas lacking these interventions. We further hypothesised that combined nutrition and sanitation interventions would lead to synergistic improvements in linear growth beyond what was realised in either standalone intervention arm. The hypothesised pathway for these effects, consistent with secondary outcome measures, was reduced enteric pathogen frequency and enteric pathogen load in stools (Figure 1).

**Figure 1:**
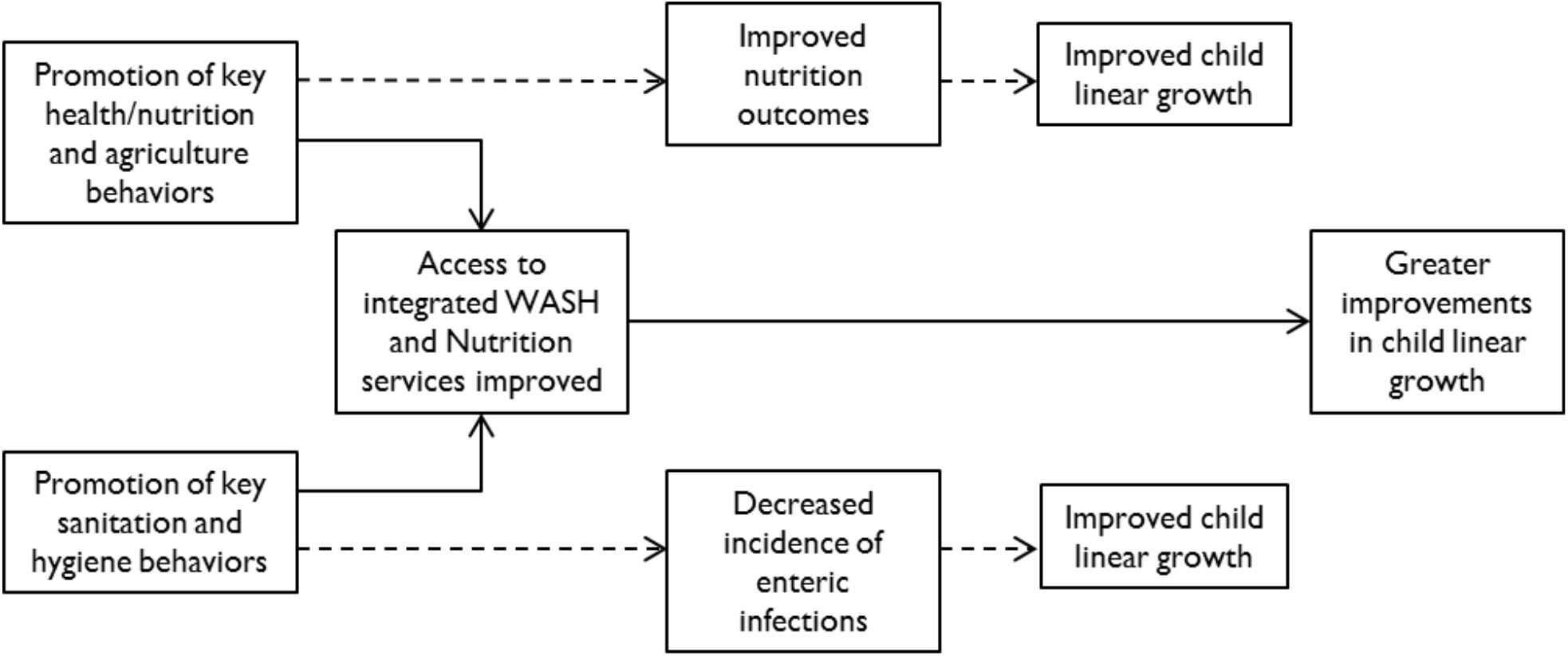
Theory of change diagram.

## Methods

### Study design and participants

We implemented a two-by-two factorial cRCT in rural communes in three provinces in Cambodia: Battambang, Pursat, and Siem Reap. The communes targeted by the program were selected based on two criteria: communes where at least 30% of the population was living below the poverty line according to the 2011 Cambodia Ministry of Planning’s Commune Database; and communes where latrine subsidies were not then in place. This study is reported per the Consolidated Standards of Reporting Trials (CONSORT) guideline (see Supplementary Material for CONSORT checklist).

### Randomisation and masking

In 2015, prior to the start of project activities, we randomly assigned communes to one of three treatment arms (nutrition only, sanitation/hygiene only, combined nutrition and sanitation/hygiene) or control arm using a random number generator with reproducible seed in Stata 13 (Stata, College Station, TX). Randomisation was conducted at the commune level to limit the risk of contamination between study arms and all villages within each commune received the assigned intervention. Following randomisation, three communes were dropped from the trial due to objections from the local governments of overlap with other current programming. This resulted in 55 communes with treatment arms of different sizes: 11 communes in nutrition-only arm; 13 in sanitation-only arm; 12 in combined-intervention arm; and 19 in control arm (Figure 2). The trial enrolled primary caregivers with a child who was born after intervention implementation began (up to 28 months prior) and who had lived in the commune during the child’s entire life, resulting in a participant population of children 1-28 months old. Neither participants nor field staff were masked to treatment status due to the nature of the interventions, but data collection teams were blinded to the arm assignment and number of treatment arms.

**Figure 2:**
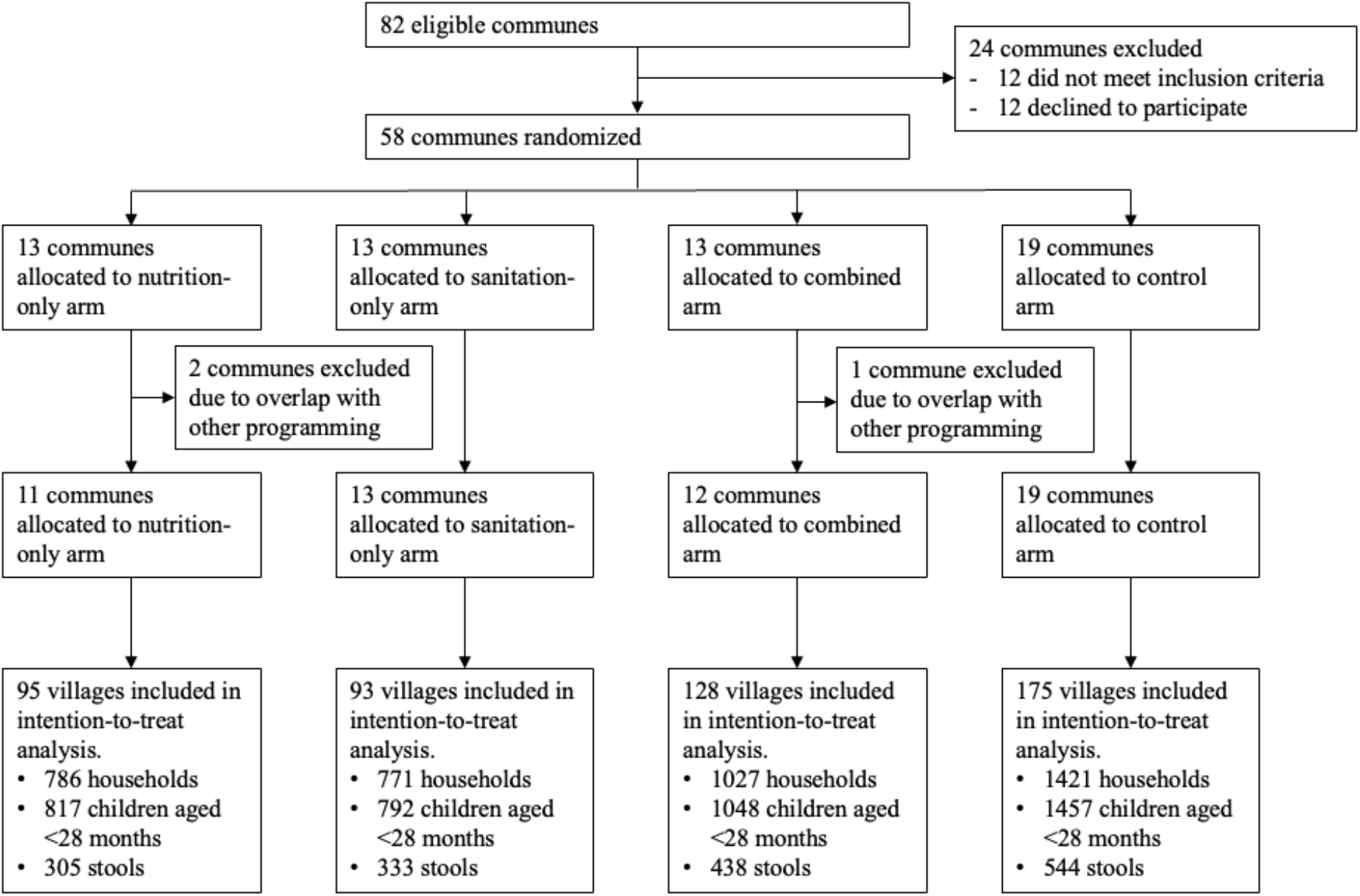
Trial profile.

### Procedures

The interventions were delivered in the 36 intervention communes over the course of two years, between 2015-2017, while the remaining 19 control communes were unexposed to the programmes. Two international non-governmental organizations—Save the Children and SNV— provided programmatic implementation and coordinated activities with local governments. The nutrition interventions included complementary feeding activities and education through community-based growth promotion sessions; caregiver groups; home visits; and conditional cash transfers (CCTs) linked to the utilization of key health and nutrition services focusing on first 1,000 days of life. The sanitation interventions consisted primarily of CLTS as it was delivered here, latrine vouchers coupled with supply-side support for sanitation and hygiene products, and social behaviours change communications (SBCC). Intervention activities and frequency are summarized in Table 1, and additional details about the interventions are described in the Supplementary Material.

**Table 1:**
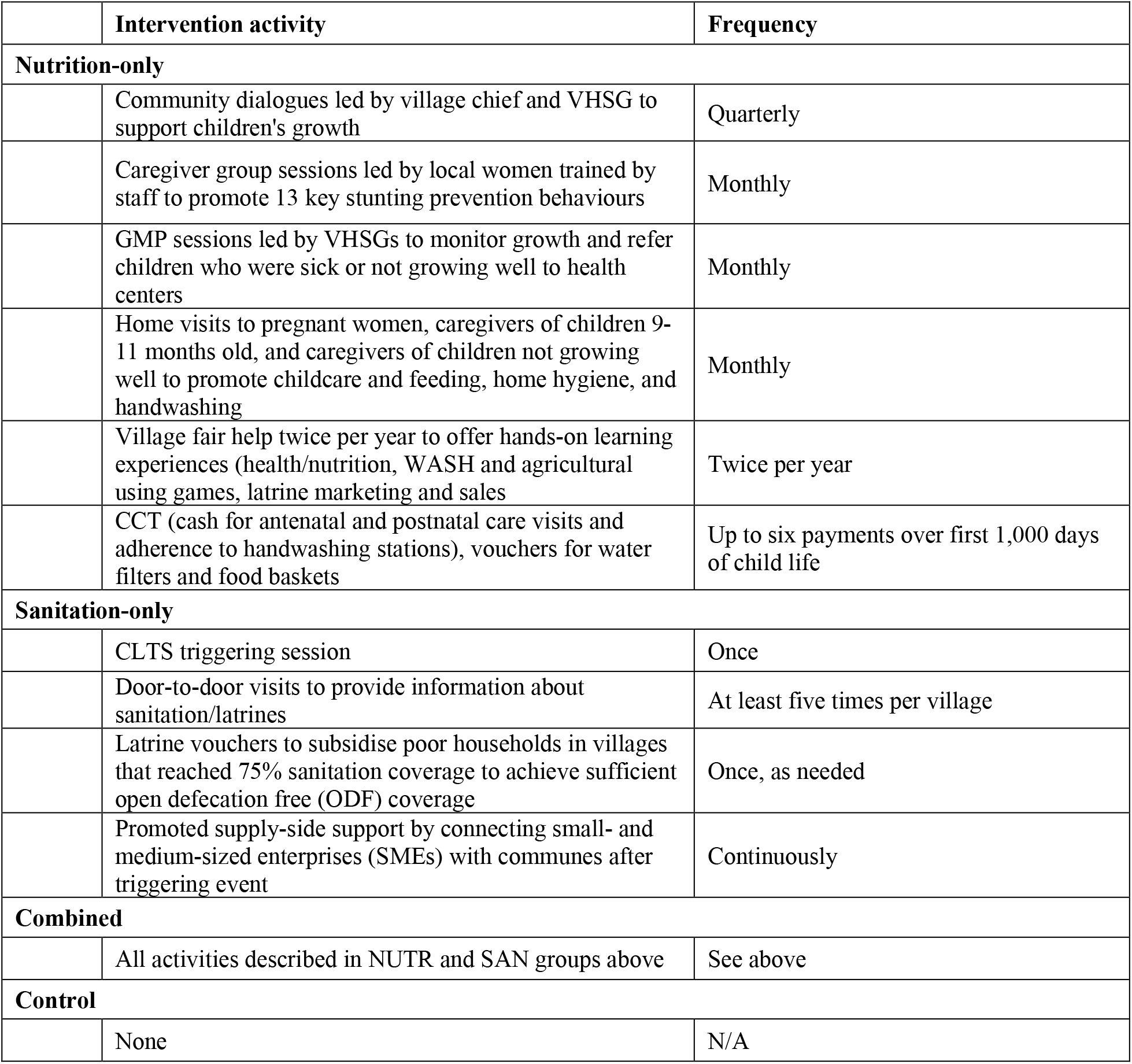
Summary of intervention activities.

|  | **Intervention activity** | **Frequency** |
| --- | --- | --- |
| **Nutrition-only** |  |  |
|  | Community dialogues led by village chief and VHSG to support children's growth | Quarterly |
|  | Caregiver group sessions led by local women trained by staff to promote 13 key stunting prevention behaviours | Monthly |
|  | GMP sessions led by VHSGs to monitor growth and refer children who were sick or not growing well to health centers | Monthly |
|  | Home visits to pregnant women, caregivers of children 9-11 months old, and caregivers of children not growing well to promote childcare and feeding, home hygiene, and handwashing | Monthly |
|  | Village fair help twice per year to offer hands-on learning experiences (health/nutrition, WASH and agricultural using games, latrine marketing and sales | Twice per year |
|  | CCT (cash for antenatal and postnatal care visits and adherence to handwashing stations), vouchers for water filters and food baskets | Up to six payments over first 1,000 days of child life |
| **Sanitation-only** |  |  |
|  | CLTS triggering session | Once |
|  | Door-to-door visits to provide information about sanitation/latrines | At least five times per village |
|  | Latrine vouchers to subsidise poor households in villages that reached 75% sanitation coverage to achieve sufficient open defecation free (ODF) coverage | Once, as needed |
|  | Promoted supply-side support by connecting small- and medium-sized enterprises (SMEs) with communes after triggering event | Continuously |
| **Combined** |  |  |
|  | All activities described in NUTR and SAN groups above | See above |
| **Control** |  |  |
|  | None | N/A |

The survey was communicated in the Khmer language to assess household and child-level risk factors of children under 28 months of age. Enumerators completed in-home interviews with the primary caregiver of children in the household about basic household member information; breastfeeding and nutrition of children up to age 28 months; number of pregnancies and child births of the caregiver; intervention exposure and participation; household WASH conditions and practices; and household assets/characteristics to construct wealth scores (excluding WASH variables). We also documented process evaluation (PE) indicators based on self-reported receipt of, and participation in, intervention activities to assess intervention fidelty and adherence, respectively. We attempted to collect a stool sample from each child and randomly selected a subset of stools for analysis by reverse-transcription quantitative polymerase chain reaction (RT-qPCR) of 30 enteric pathogen genes using a custom-developed TaqMan Array Card (TAC; ThermoFisher Scientific, Waltham, VA), as described in the Supplementary Material.

### Outcomes

The primary outcome was length-for-age Z-score (LAZ). For children 1-24 months in age, we measured recumbent length; for children 24-28 months in age, we measured standing height. Herein, “length” will be inclusive of both recumbent length and standing height. Secondary outcomes included weight-for-age Z-score (WAZ); weight-for-length Z-score (WLZ); proportion of children stunted (LAZ<-2), underweight (WAZ<-2), and wasted (WLZ<-2); caregiver reported diarrhoea; all-cause mortality; and enteric pathogen detection and quantification in stool. Child length and weight were measured by trained paired enumerators following guidelines from the National Health and Nutrition Examination Survey (NHANES)^16^. Final measurements took place in August 2019, 28 months after the end of the roll-out period. Data collection was completed by KHANA Centre for Population Health Research, with oversight and support from Management Systems International (MSI). Data collection details, measurement protocols, and PE indicators are further described in Supplementary Material.

We assessed enteric pathogens as the prevalence of individual gene targets, the number of co-detected pathogens, and enteric pathogen-associated gene copies per gram of stool based on PCR quantification cycle (Cq) and standard curves. *E.coli* pathotypes were defined as: EAEC (*aaiC*, or *aatA*, or both), atypical EPEC (*eae* without *bfpA, stx1*, and *stx2*), typical EPEC (*bfpA*), ETEC (*STh, STp*, or *LT*), and STEC (*eae* without *bfpA* and with *stx1, stx2*, or both). Details on nucleic acid extraction and molecular assaying are described in Supplementary Material.

### Statistical analysis

We performed an intention-to-treat (ITT) analysis for all outcomes using generalised estimating equations (GEE) with robust standard errors to account for clustering at the village level. We did not consider pre-intervention covariate balance^17^ but present secondary analyses adjusted for pre-specified pre-intervention covariates in the Supplementary Material. Outcomes in each treatment arm were compared to the control arm and between standalone treatment arms and the combined treatment arm. We used linear regression to estimate mean differences in LAZ, WAZ, WLZ, and log_10_-transformed pathogen gene target densities and used log-linear Poisson regression to estimate the prevalence ratio (PR) between arms for nutritional status (stunting, wasting, and underweight), diarrhoea, and overall mortality. Enteric pathogen gene outcomes were dichotomised, with positive detections defined by a Cq <35^18^, and Poisson regression was used to estimate PRs for individual pathogens detected in stool. We further estimated the incidence rate ratio (IRR) of co-detected pathogens (total and in subgroups by bacteria, viruses, protozoa, and STHs) using negative binomial regression. We did not adjust for multiple comparisons for growth, diarrhoea, or mortality outcomes^3,19^, but we did apply the Benjamini-Hochberg procedure to control the false discovery rate within analyses of multiple enteric pathogen outcomes^20^. Details on power calculations are included in Supplementary Material.

### Ethics

The study received approval from the National Ethics Committee for Health Research in the Cambodian Ministry of Health, Georgia Institute of Technology, and New England Institutional Review Board. Prior to any data collection, the trial was explained to participants in the Khmer language. Written and verbal consent were obtained prior to administering the surveys and anthropometry measurements. The trial was pre-registered with ISRCTN Registry (<u>ISRCTN77820875</u>).

## Results

Among 82 presumptively eligible communes, the provincial governments in 27 declined to participate. Ultimately, the evaluation included 55 communes randomly assigned to one of three treatment arms (n=36 communes) or control arm (n=19 communes); the control arm was relatively oversized to enhance statistical efficiency of multiple hypothesis testing^21^. Figure 2 shows the trial profile by intervention subgroups. 4,015 households participated in endline surveys; 4,005 households were included in these analyses (10 were excluded due to incomplete surveys), and 4,124 children had anthropometry measures taken.

Household and caregiver characteristics were mostly similar across treatment and control groups (Table 2). Primary caregivers in the control group reported lower levels of primary school attendance compared to the treatment groups, but paternal primary school attendance was similar. Households in the nutrition-only and sanitation-only groups had higher wealth index scores compared to households in the combined intervention and control groups. The control group had a higher prevalence of improved water source as their main source of drinking water compared to the treatment groups.

**Table 2:**
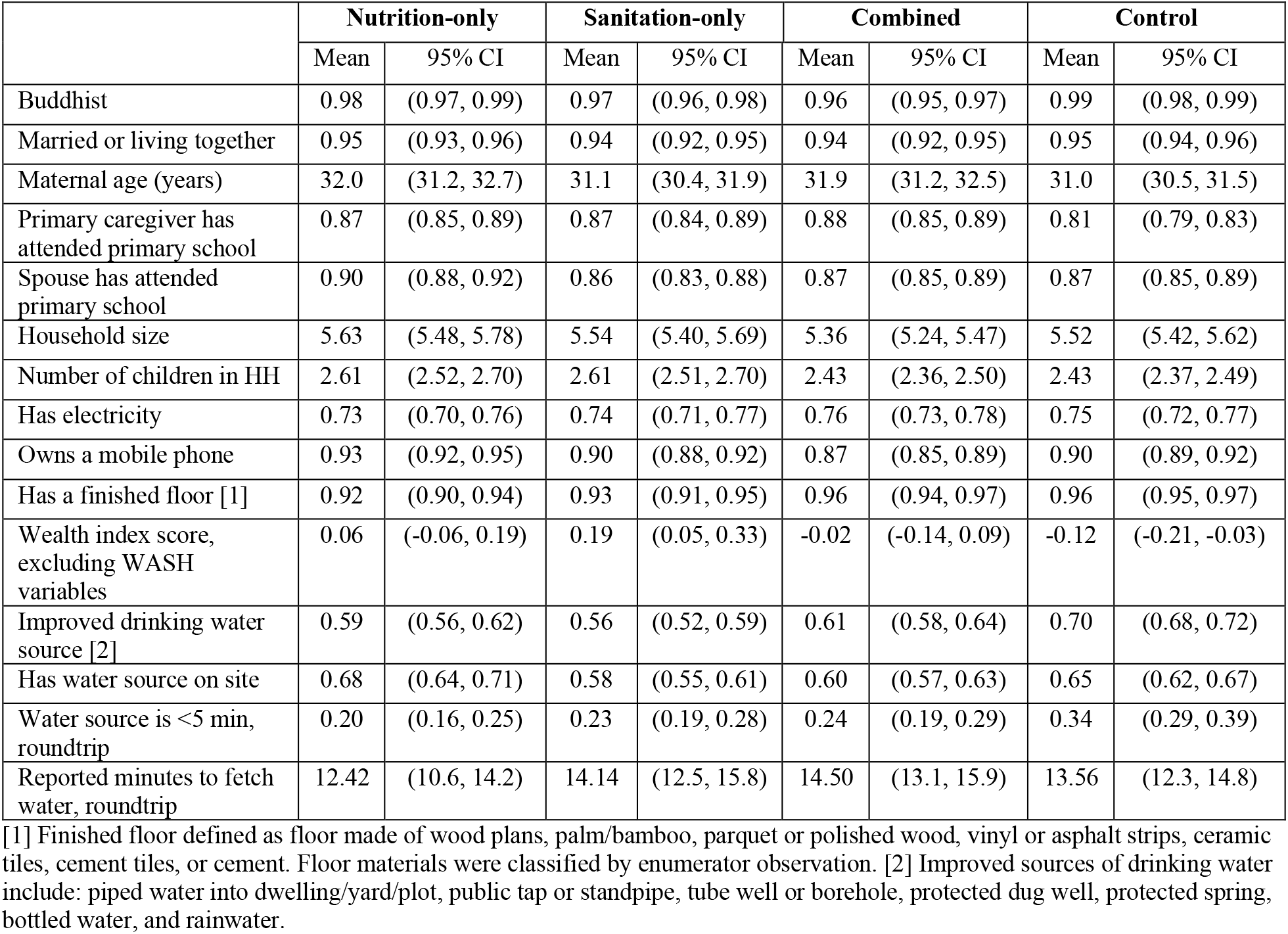
Household and caregiver characteristics.

Nutrition intervention fidelity was high, with households in the nutrition-only and combined-intervention arms reporting significantly higher participation in these activities compared to the sanitation-only and control groups (Table 3). Approximately 60% of households in the nutrition-only and combined-intervention arms reported participating in at least four of the eight nutrition intervention activities, compared to 4% in the sanitation-only and control arms. Conversely, sanitation intervention fidelity was very low, with only 6% of households in the sanitation-only and control arms reporting participation in any CLTS activity, compared to 14% of households in the nutrition-only arm and 25% in the combined-intervention arm.

**Table 3:** Intervention fidelity indicators.

|  | Nutrition-only |  |  | Sanitation-only |  |  | Combined |  |  | Control |  |  |
| --- | --- | --- | --- | --- | --- | --- | --- | --- | --- | --- | --- | --- |
| **Nutrition** | N | n | % | N | n | % | N | n | % | N | n | % |
| Participated in any "First 1,000 Days" type activity [1] | 817 | 615 | 75% | 792 | 145 | 18% | 1,055 | 813 | 77% | 1,460 | 383 | 26% |
| Participated in any GMP | 817 | 641 | 78% | 792 | 181 | 23% | 1,055 | 935 | 89% | 1,460 | 482 | 33% |
| Received home visit VHSG | 817 | 518 | 63% | 792 | 227 | 29% | 1,055 | 661 | 63% | 1,460 | 490 | 34% |
| Enrolled in any CCT program for health and nutrition [2] | 817 | 224 | 27% | 792 | 19 | 2% | 1,055 | 228 | 22% | 1,460 | 31 | 2% |
| Received any voucher for food basket [3] | 817 | 440 | 54% | 792 | 1 | 0% | 1,055 | 554 | 53% | 1,460 | 6 | 0% |
| Received any voucher for water filter [3] | 817 | 41 | 5% | 792 | 40 | 5% | 1,055 | 149 | 14% | 1,460 | 100 | 7% |
| Aware of *Grow Together* campaign [4] | 817 | 353 | 43% | 792 | 93 | 12% | 1,055 | 471 | 45% | 1,460 | 149 | 10% |
| Participation in nutrition intervention activities: none (0 of 8) | 817 | 65 | 8% | 792 | 404 | 51% | 1,055 | 44 | 4% | 1,460 | 585 | 40% |
| Participation in nutrition intervention activities: low (1-3 of 8) | 817 | 262 | 32% | 792 | 359 | 45% | 1,055 | 367 | 35% | 1,460 | 809 | 55% |
| Participation in nutrition intervention activities: med (4-6 of 8) | 817 | 387 | 47% | 792 | 29 | 4% | 1,055 | 501 | 47% | 1,460 | 65 | 4% |
| Participation in nutrition intervention activities: high (7-8 of 8) | 817 | 103 | 13% | 792 | 0 | 0% | 1,055 | 143 | 14% | 1,460 | 1 | 0% |
| **Sanitation** |  |  |  |  |  |  |  |  |  |  |  |  |
| Any CLTS participation [5] | 817 | 115 | 14% | 792 | 46 | 6% | 1,055 | 261 | 25% | 1,460 | 81 | 6% |
| Received any voucher to build latrine [6] | 817 | 66 | 8% | 792 | 51 | 6% | 1,055 | 123 | 12% | 1,460 | 84 | 6% |
| [1] "First 1,000 Days" activities were administered in nutrition-only and combined arms and include: community dialogues, caregiver group education sessions, and village fairs. [2] CCT program ended in Jan 2019 and the Government of Cambodia started a new CCT program in July 2019 across study area. CCT program administered in nutrition-only and combined arms only. [3] Vouchers for water filter and food baskets were targeted subsidies distributed to CCT participants in nutrition-only and combined arms. [4] *Grow Together* campaign was part of the nutrition programming (nutrition-only and combined arms). However, three TV spots were seen across all four arms. [5] In sanitation-only and combined arms, the Ministry of Rural Development confirmed that the project was the only CLTS campaign active in those areas. [6] Latrine vouchers were targeted subsidies given to households in villages that reached 75% sanitation coverage in sanitation-only and combined arms. |  |  |  |  |  |  |  |  |  |  |  |  |

More households in the control arm (70%) had an improved water source as their main source of drinking water, an indicator of nutrition intervention adherence, compared to other arms (approximately 60% in other arms; Table 4). The combined intervention arm had greater access to improved sanitation facilities (61%) compared to the nutrition-only (55%), sanitation-only (51%), and control (52%) arms. OD (self-reported) was practiced less in the combined intervention arm (7%) compared to the nutrition-only (14%), sanitation-only (16%), and control (16%) arms. Notably, the sanitation-only arm experienced a significantly larger increase in sanitation coverage (+25 percentage points [pp]) compared to all other arms (+14pp in nutrition-only arm, +19pp in combined and control arms), though sanitation gains across all arms were evident in the intervention period, reflecting a strong secular trend of sanitation expansion that has been widely documented in rural Cambodia^8,10,22,23^. Additional intervention adherence indicators related to environmental hygiene are reported in the Supplementary Material.

**Table 4:** Intervention adherence indicators (28-months after intervention)

|  | Nutrition-only |  |  | Sanitation-only |  |  | Combined |  |  | Control |  |  |
| --- | --- | --- | --- | --- | --- | --- | --- | --- | --- | --- | --- | --- |
| **Nutrition** | N | n or mean | % or SD | N | n or mean | % or SD | N | n or mean | % or SD | N | n or mean | % or SD |
| Visited health facility for at least four antenatal care check-ups | 697 | 632 | 91% | 712 | 646 | 91% | 910 | 819 | 90% | 1,257 | 1116 | 89% |
| Brought child for monthly GMP at community or health center | 817 | 641 | 78% | 792 | 181 | 23% | 1,055 | 935 | 89% | 1,460 | 482 | 33% |
| Breastfeeding exclusively for children <6 months | 171 | 109 | 64% | 161 | 110 | 68% | 205 | 140 | 68% | 272 | 182 | 67% |
| Ever breastfed (all children) | 817 | 797 | 98% | 792 | 765 | 97% | 1,048 | 1021 | 97% | 1,457 | 1420 | 97% |
| Solid and semi-solid foods eaten for children >6 months | 646 | 609 | 94% | 631 | 611 | 97% | 843 | 792 | 94% | 1,185 | 1134 | 96% |
| Dietary diversity score (0-7) | 817 | 2.24 | 1.61 | 792 | 2.19 | 1.58 | 1,048 | 2.20 | 1.62 | 1,457 | 2.33 | 1.59 |
| Achieved minimum dietary diversity | 817 | 202 | 25% | 792 | 177 | 22% | 1,048 | 247 | 24% | 1,457 | 350 | 24% |
| Achieved minimum meal frequency | 817 | 537 | 66% | 792 | 518 | 65% | 1,048 | 680 | 65% | 1,457 | 977 | 67% |
| Achieved minimum acceptable diet | 817 | 170 | 21% | 792 | 159 | 20% | 1,048 | 205 | 20% | 1,457 | 310 | 21% |
| Treated drinking water | 817 | 548 | 67% | 792 | 471 | 59% | 1,055 | 765 | 73% | 1,460 | 1,037 | 71% |
| Treated drinking water with filter | 817 | 151 | 18% | 792 | 160 | 20% | 1,055 | 302 | 29% | 1,460 | 548 | 40% |
| **Sanitation** |  |  |  |  |  |  |  |  |  |  |  |  |
| Had improved sanitation facility [1] | 816 | 452 | 55% | 791 | 400 | 51% | 1,054 | 638 | 61% | 1,459 | 759 | 52% |
| Open defecation (OD) | 817 | 112 | 14% | 792 | 126 | 16% | 1,055 | 73 | 7% | 1,460 | 231 | 16% |
| Used shared toilet | 817 | 252 | 31% | 791 | 264 | 33% | 1,054 | 343 | 33% | 1,459 | 468 | 32% |
| Caregiver reported adults in HH openly defecating | 697 | 92 | 13% | 658 | 116 | 18% | 973 | 118 | 12% | 1,208 | 213 | 18% |
| Time to get to toilet, one way (minutes) | 171 | 4.22 | 4.11 | 166 | 3.92 | 3.83 | 219 | 4.74 | 8.17 | 291 | 5.05 | 8.27 |
| Reported latrine built as a result of CLTS activity | 115 | 51 | 44% | 46 | 15 | 33% | 261 | 91 | 35% | 81 | 28 | 35% |
| Reported latrine built using latrine voucher | 50 | 10 | 20% | 15 | 4 | 27% | 91 | 37 | 41% | 28 | 12 | 43% |
| Main reason for not constructing latrine: lack of funds | 20 | 17 | 85% | 18 | 14 | 78% | 60 | 55 | 92% | 14 | 14 | 100% |
| Main reason for construction latrine: privacy | 51 | 7 | 14% | 15 | 6 | 40% | 91 | 6 | 7% | 28 | 7 | 25% |
| Main reason for construction latrine: security | 51 | 10 | 20% | 15 | 2 | 13% | 91 | 20 | 22% | 28 | 4 | 14% |
| Main reason for construction latrine: hygiene | 51 | 17 | 33% | 15 | 5 | 33% | 91 | 43 | 47% | 28 | 10 | 36% |
| Main reason for construction latrine: OD is harmful | 51 | 5 | 10% | 15 | 1 | 7% | 91 | 9 | 10% | 28 | 5 | 18% |
| Child stools properly disposed of [2] | 817 | 581 | 71% | 792 | 515 | 65% | 1,055 | 781 | 74% | 1,460 | 993 | 68% |
| Community-level open defecation before intervention | 817 | 28% | 27% | 792 | 41% | 33% | 1,055 | 28% | 28% | 1,460 | 32% | 31% |
| Community-level open defecation after intervention | 817 | 14% | 21% | 792 | 16% | 21% | 1,055 | 9% | 17% | 1,460 | 13% | 20% |
| **Environmental hygiene** |  |  |  |  |  |  |  |  |  |  |  |  |
| Child stools left in the open | 817 | 147 | 18% | 792 | 170 | 21% | 1,055 | 160 | 15% | 1,460 | 315 | 22% |
| Child play environment observed to be free of animals | 817 | 182 | 22% | 792 | 187 | 24% | 1,055 | 261 | 25% | 1,460 | 294 | 20% |
| Child play environment observed to be free of garbage/HH waste | 817 | 298 | 36% | 792 | 290 | 37% | 1,055 | 419 | 40% | 1,460 | 567 | 39% |
| Child play environment observed to be free of sharp objects | 817 | 449 | 55% | 792 | 427 | 54% | 1,055 | 639 | 61% | 1,460 | 818 | 56% |
| Child play environment observed to be free of faeces | 817 | 313 | 38% | 792 | 304 | 38% | 1,055 | 448 | 42% | 1,460 | 555 | 38% |
| [1] Improved sanitation facilities include: flush/pour flush toilet to a piped sewer system, septic tank or pit latrine, a ventilated improved pit latrine, a pit latrine with slab, and a composting toilet. [2] Proper disposal of children faeces consist of putting or rinsing stool into a sanitation facility or burying it; improper disposal of children faeces includes putting or rinsing stool into a drain or ditch, throwing it into garbage or leaving it in the open. |  |  |  |  |  |  |  |  |  |  |  |  |

### Primary and secondary outcomes

Mean LAZ in the control arm was -1.04 (SD 1.20). Compared with control, children in the nutrition-only arm were longer by a mean of 0.08 LAZ (95% CI -0.01, 0.18), and children in the combined-intervention arm were longer by 0.10 LAZ (95% CI 0.01, 0.20), although these differences were not observed in the adjusted analyses (

Table 5, Figure 3, Supplementary Material). Children in the nutrition-only arm and combined-intervention arm were heavier than children in the control arm by a mean of 0.10 WAZ (95% CI 0.00, 0.19) and 0.11 (95% CI 0.03, 0.20), respectively. These differences were slightly attenuated in the adjusted analyses (Table 5, Supplementary Material). No differences were observed between the control arm and intervention arms in terms of WLZ. Children in the combined intervention arm were also longer and heavier, on average, than children in the sanitation-only arm by 0.16 LAZ (95% CI 0.04, 0.27) and 0.10 WAZ (95% CI 0.01, 0.20), respectively. LAZ and WAZ were similar between children in the nutrition-only and combined intervention arms.

**Table 5:** Effects of interventions on length and weight (Primary outcome (LAZ) and secondary outcomes (WAZ, WLZ)), comparing intervention arms to control and single intervention arms to combined intervention.

|  |  |  |  | Unadjusted mean difference (95% CI) |  |
| --- | --- | --- | --- | --- | --- |
|  | N | Mean | SD | Compared to control arm | Compared to combined intervention arm |
| LAZ |  |  |  |  |  |
| Nutrition-only | 798 | -0.95 | 1.16 | 0.08 (-0.01, 0.18) | -0.02 (-0.12, 0.09) |
| Sanitation-only | 777 | -1.09 | 1.23 | -0.05 (-0.16, 0.05) | -0.16 (-0.27, -0.04) |
| Combined | 1037 | -0.94 | 1.16 | 0.10 (0.01, 0.20) | -- |
| Control | 1443 | -1.04 | 1.20 | -- | -- |
| WAZ |  |  |  |  |  |
| Nutrition-only | 815 | -0.95 | 1.29 | 0.10 (0.00, 0.19) | -0.02 (-0.12, 0.08) |
| Sanitation-only | 792 | -1.04 | 1.13 | 0.01 (-0.07, 0.09) | -0.10 (-0.20, -0.01) |
| Combined | 1044 | -0.94 | 1.11 | 0.11 (0.03, 0.20) | -- |
| Control | 1452 | -1.05 | 1.10 | -- | -- |
| WLZ |  |  |  |  |  |
| Nutrition-only | 814 | -0.60 | 1.04 | 0.06 (-0.03, 0.15) | -0.02 (-0.12, 0.08) |
| Sanitation-only | 790 | -0.59 | 0.98 | 0.06 (-0.02, 0.14) | -0.02 (-0.11, 0.07) |
| Combined | 1043 | -0.58 | 1.03 | 0.08 (0.00, 0.16) | -- |
| Control | 1452 | -0.65 | 0.98 | -- | -- |

**Figure 3:**
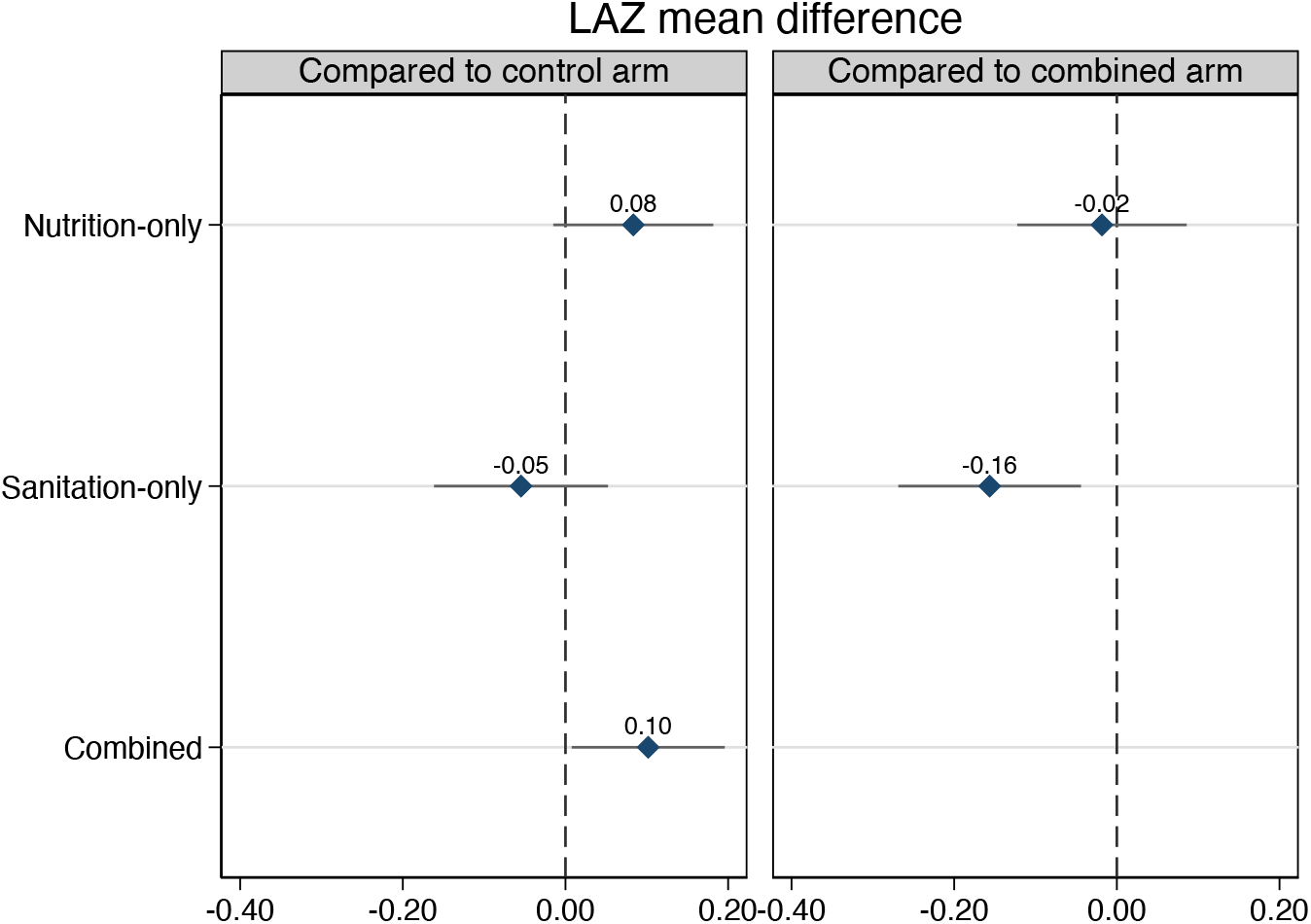
Unadjusted intervention effects on LAZ. Estimates are mean differences (point) with 95% CIs (line)

**Figure 4:**
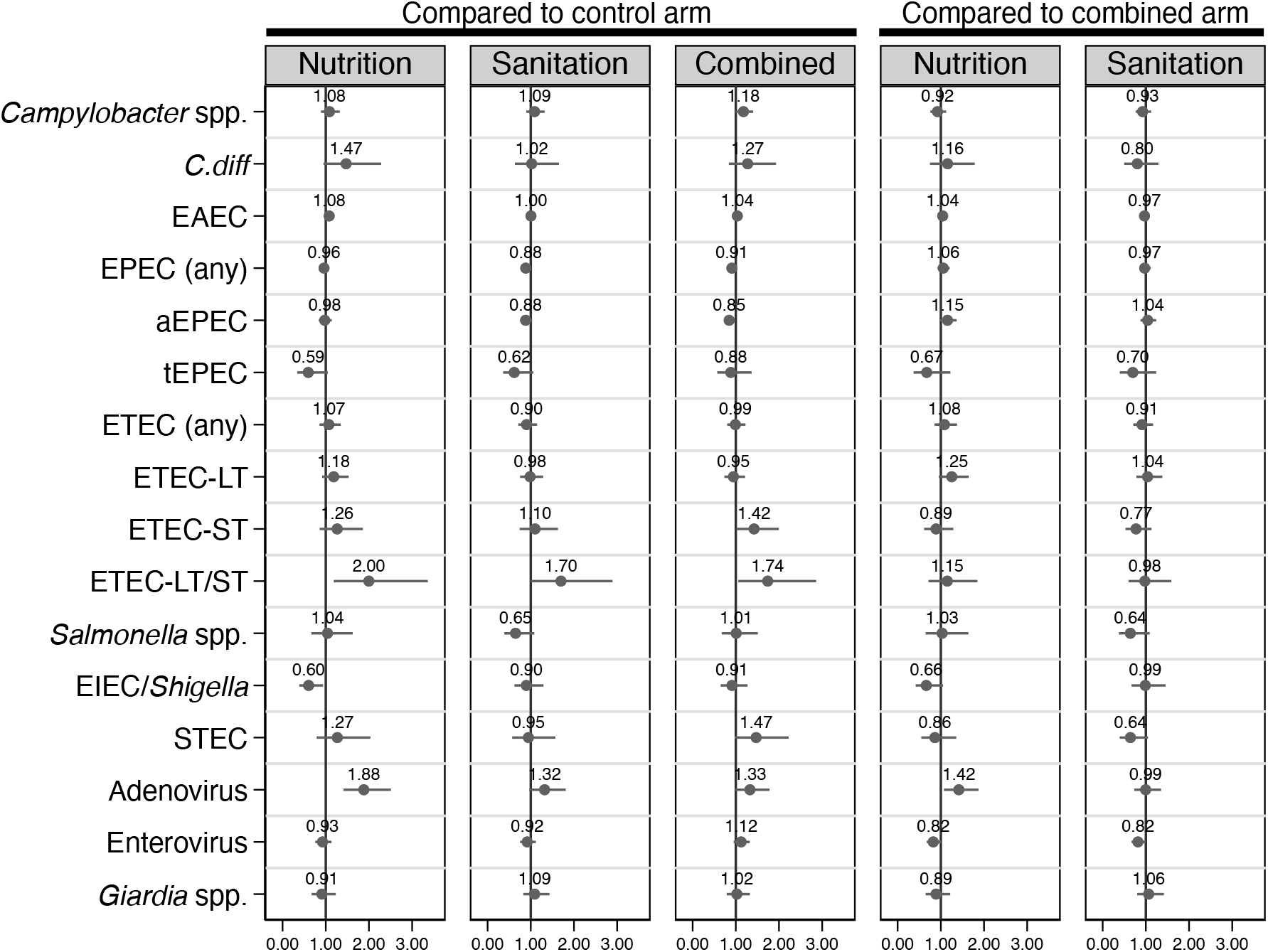
Impact of interventions on unadjusted prevalence ratio of individual pathogens. Point estimates and 95% confidence intervals were determined using log-linear Poisson models with generalized estimating equations.

Compared with the control arm, none of the intervention arms differed in the prevalence of children who experienced stunting, wasting, diarrhoea (7-day recall), or mortality (Table 6). However, the combined intervention reduced underweight prevalence by 18% (PR 0.82, 95% CI 0.68, 0.99) relative to the control arm. Although the combined intervention did not significantly impact stunting prevalence compared with the control arm or the nutrition-only arm, the sanitation-only arm was associated with a 20% increase (PR 1.2, 95% CI 1.0, 1.5) in the prevalence of both stunting and underweight status when compared to the combined intervention. All associations with stunting and underweight were attenuated in adjusted analyses (Supplementary Material).

**Table 6:** Effects of intervention on child health outcomes, comparing intervention arms to control and single intervention arms to combined intervention.

|  |  |  |  | Compared to control arm | Compared to combined-intervention arm |
| --- | --- | --- | --- | --- | --- |
|  | N | Mean | SD | PR (95% CI) | PR (95% CI) |
| **Stunted** |  |  |  |  |  |
| Nutrition-only | 801 | 0.15 | 0.36 | 0.84 (0.69, 1.03) | 0.93 (0.74, 1.15) |
| Sanitation-only | 782 | 0.21 | 0.40 | 1.12 (0.94, 1.33) | 1.23 (1.02, 1.49) |
| Combined | 1046 | 0.17 | 0.37 | 0.91 (0.76, 1.09) | -- |
| Control | 1449 | 0.18 | 0.39 | -- | -- |
| **Wasted** |  |  |  |  |  |
| Nutrition-only | 815 | 0.07 | 0.26 | 0.87 (0.65, 1.17) | 1.12 (0.80, 1.57) |
| Sanitation-only | 790 | 0.07 | 0.26 | 0.84 (0.62, 1.14) | 1.08 (0.76, 1.53) |
| Combined | 1052 | 0.07 | 0.25 | 0.78 (0.58, 1.04) | -- |
| Control | 1457 | 0.08 | 0.28 | -- | -- |
| **Underweight** |  |  |  |  |  |
| Nutrition-only | 816 | 0.15 | 0.35 | 0.85 (0.71, 1.03) | 1.04 (0.84, 1.29) |
| Sanitation-only | 792 | 0.17 | 0.38 | 1.00 (0.85, 1.19) | 1.22 (1.00, 1.49) |
| Combined | 1053 | 0.14 | 0.35 | 0.82 (0.68, 0.99) | -- |
| Control | 1457 | 0.17 | 0.38 | -- | -- |
| **Diarrhoea (7-day recall)** |  |  |  |  |  |
| Nutrition-only | 788 | 0.19 | 0.39 | 0.89 (0.74, 1.06) | 0.95 (0.78, 1.14) |
| Sanitation-only | 752 | 0.21 | 0.41 | 0.99 (0.84, 1.17) | 1.05 (0.88, 1.25) |
| Combined | 1018 | 0.20 | 0.40 | 0.94 (0.80, 1.11) | -- |
| Control | 1411 | 0.21 | 0.41 | -- | -- |
| **All-cause mortality** |  |  |  |  |  |
| Nutrition-only | 1574 | 0.03 | 0.16 | 1.55 (0.71, 3.39) | 1.61 (0.68, 3.82) |
| Sanitation-only | 1636 | 0.03 | 0.16 | 1.09 (0.50, 2.40) | 1.13 (0.48, 2.68) |
| Combined | 1932 | 0.03 | 0.16 | 0.96 (0.44, 2.10) | -- |
| Control | 2688 | 0.03 | 0.16 | -- | -- |

### Enteric pathogen results

We assessed enteric pathogen-associated gene targets in 1,620 randomly selected stools that demonstrated acceptible amplification (of 4,114 stools total, see Supplementary Material): 305 from the nutrition arm, 333 from the sanitation arm, 438 from the combined-intervention arm, and 544 from the control arm. We detected at least one bacterial gene in 87% of all samples, at least one viral gene in 49% of samples, at least one protozoan gene in 20% of samples, and at least one STH gene in 2% of samples. Enteroaggregative *E. coli* (EAEC), enteric pathogenic *E. coli* (EPEC), enterovirus, *Campylobacter* spp., and enterotoxigenic *E. coli* (ETEC) were the most prevalent pathogens (Table 7). We detected a mean 2.2 bacterial genes (out of 9), 0.59 viral genes (out of 6), 0.21 protozoan genes (out of 4), and 0.03 STH genes (out of 4) in each sample. We found no differences in the rate of bacterial, viral, protozoan, or STH gene co-detection between the control arm and any treatment arm or between the combined arm and the standalone intervention arms (Table 8). Prevalence increased with age for many pathogens (aEPEC, ETEC, *Shigella*/EIEC (*ipah*), STEC, adenovirus, *Giardia*), while prevalence peaked for children 9-17 months for other pathogens (*Campylobacter* spp., *C.diff*, EAEC, *Salmonella* spp.; Supplementary Material).

**Table 7:** Enteric pathogen gene prevalence among treatment arms.

|  | All samples<br>(N=1620) | Nutrition-only<br>(N=305) | Sanitation-only<br>(N=333) | Combined<br>(N=438) | Control<br>(N=544) |
| --- | --- | --- | --- | --- | --- |
| **Bacteria** |  |  |  |  |  |
| *Campylobacter* spp. | 551 (34%) | 104 (34%) | 114 (34%) | 162 (37%) | 171 (31%) |
| *Clostridium difficile* | 139 (9%) | 33 (11%) | 25 (8%) | 41 (9%) | 40 (7%) |
| EAEC | 1029 (64%) | 204 (67%) | 207 (62%) | 281 (64%) | 337 (62%) |
| EPEC | 899 (55%) | 172 (56%) | 173 (52%) | 234 (53%) | 320 (59%) |
| aEPEC | 703 (43%) | 139 (46%) | 137 (41%) | 173 (39%) | 254 (47%) |
| tEPEC | 109 (7%) | 15 (5%) | 17 (5%) | 32 (7%) | 45 (8%) |
| ETEC | 422 (26%) | 86 (28%) | 79 (24%) | 114 (26%) | 143 (26%) |
| ETEC-LT | 342 (21%) | 75 (25%) | 68 (20%) | 86 (20%) | 113 (21%) |
| ETEC-ST | 194 (12%) | 39 (13%) | 37 (11%) | 63 (14%) | 55 (10%) |
| ETEC-LT/ST | 114 (7%) | 28 (9%) | 26 (8%) | 35 (8%) | 25 (5%) |
| *Salmonella* spp. | 134 (8%) | 28 (9%) | 19 (6%) | 39 (9%) | 48 (9%) |
| *Shigella* spp. | 186 (11%) | 24 (8%) | 39 (12%) | 52 (12%) | 71 (13%) |
| STEC | 132 (8%) | 27 (9%) | 22 (7%) | 45 (10%) | 38 (7%) |
| *Vibrio cholerae* | 10 (1%) | 1 (0%) | 6 (2%) | 1 (0%) | 2 (0%) |
| At least 1 bacterium detected | 1410 (87%) | 276 (90%) | 282 (85%) | 386 (88%) | 466 (86%) |
| Mean number of bacteria detected | 2.48<br>(2.42, 2.55) | 2.46<br>(2.32, 2.60) | 2.43<br>(2.28, 2.57) | 2.51<br>(2.39, 2.63) | 2.51<br>(2.40, 2.62) |
| **Viruses** |  |  |  |  |  |
| Adenovirus | 287 (18%) | 77 (25%) | 59 (18%) | 78 (18%) | 73 (13%) |
| Astrovirus | 7 (0%) | 2 (1%) | 1 (0%) | 3 (1%) | 1 (0%) |
| Enterovirus | 558 (34%) | 97 (32%) | 105 (32%) | 169 (39%) | 187 (34%) |
| Norovirus | 54 (3%) | 8 (3%) | 9 (3%) | 20 (5%) | 17 (3%) |
| Rotavirus | 17 (1%) | 2 (1%) | 2 (1%) | 6 (1%) | 7 (1%) |
| Sapovirus | 24 (1%) | 4 (1%) | 5 (2%) | 9 (2%) | 6 (1%) |
| At least 1 virus detected | 788 (49%) | 152 (50%) | 157 (47%) | 231 (53%) | 248 (46%) |
| Mean number of viruses detected | 1.20<br>(1.17, 1.23) | 1.25<br>(1.17, 1.33) | 1.15<br>(1.09, 1.21) | 1.23<br>(1.17, 1.30) | 1.17<br>(1.12, 1.23) |
| **Protozoa** |  |  |  |  |  |
| *Cryptosporidium* | 17 (1%) | 4 (1%) | 1 (0%) | 6 (1%) | 6 (1%) |
| *Entamoeba* | 13 (1%) | 1 (0%) | 1 (0%) | 8 (2%) | 3 (1%) |
| *Giardia* | 306 (19%) | 52 (17%) | 68 (20%) | 84 (19%) | 102 (19%) |
| At least 1 protozoan detected | 328 (20%) | 56 (18%) | 69 (21%) | 93 (21%) | 110 (20%) |
| Mean number of protozoa detected | 1.02<br>(1.01, 1.04) | 1.02<br>(0.98, 1.05) | 1.01<br>(0.99, 1.04) | 1.05<br>(1.01, 1.10) | 1.01<br>(0.99, 1.03) |
| **STH** |  |  |  |  |  |
| *Ascaris lumbricoides* | 3 (0%) | 0 (0%) | 0 (0%) | 3 (1%) | 0 (0%) |
| *Trichuris trichiura* | 3 (0%) | 1 (0%) | 1 (0%) | 0 (0%) | 1 (0%) |
| *Ancylostoma duodenale* | 17 (1%) | 0 (0%) | 4 (1%) | 4 (1%) | 9 (2%) |
| *Necator americanus* | 20 (1%) | 3 (1%) | 6 (2%) | 5 (1%) | 6 (1%) |
| At least 1 | 37 (2%) | 4 (1%) | 10 (3%) | 9 (2%) | 14 (3%) |
| Mean number of STH detected | 1.16<br>(1.04, 1.29) | 1.00<br>(1.00, 1.00) | 1.10<br>(0.90, 1.30) | 1.33<br>(1.00, 1.67) | 1.14<br>(0.95, 1.34) |

**Table 8:** Unadjusted incidence rate ratios of co-detected bacteria, viruses, protozoa, and STHs, comparing intervention arms to control and single intervention arms to combined intervention.

|  | N | Compared to control arm |  |  | Compared to combined arm |  |
| --- | --- | --- | --- | --- | --- | --- |
|  |  | Nutrition-only | Sanitation-only | Combined | Nutrition-only | Sanitation-only |
| Bacteria | 1620 | 1.04<br>(0.95, 1.13) | 0.96<br>(0.87, 1.05) | 1.03<br>(0.95, 1.12) | 0.01<br>(-0.08, 0.10) | -0.07<br>(-0.17, 0.02) |
| Viruses | 1620 | 1.16<br>(0.99, 1.37) | 1.02<br>(0.86, 1.19) | 1.22<br>(1.05, 1.41) | -0.04<br>(-0.21, 0.12) | -0.18<br>(-0.34, -0.02) |
| Protozoa | 1620 | 0.92<br>(0.68, 1.23) | 1.03<br>(0.79, 1.35) | 1.10<br>(0.85, 1.41) | -0.18<br>(-0.48, 0.12) | -0.06<br>(0.66, -0.34) |
| STHs were omitted because <5% of samples had detectable STH genes. |  |  |  |  |  |  |

Examining prevalence of specific targets compared to the control arm, the nutrition-only arm demonstrated increased prevalence of any bacterial gene (PR 1.06, 95% CI 1.01, 1.11), adenovirus (PR 1.88, 95% CI 1.41, 2.51) and heat-labile/heat-stable ETEC (PR 2.00, 95% CI 1.19, 3.36) and reduced prevalence of EIEC/*Shigella* spp. (PR 0.60, 95% CI 0.39, 0.94). Children in the sanitation-only arm had less EPEC (PR 0.88, 95% CI 0.78, 1.00) compared to control. In the combined-intervention arm, atypical-EPEC prevalence decreased (PR 0.85, 95% CI 0.73, 0.98) while the prevalence increased for heat-stable ETEC (PR 1.42, 95% CI 1.01, 2.00), heat-labile/heat-stable ETEC (PR 1.74, 95% CI 1.06, 2.86), and any viral gene (PR 1.16, 95% CI 1.02, 1.31). We found similar mixed effects when comparing pathogen gene prevalence in individual treatment arms compared to the combined arm; there was slightly lower combined prevalence of any bacterial target (PR 0.96, 95% CI 0.91, 1.02) and enterovirus (PR 0.82, 95% CI 0.67, 1.00) in the sanitation-only arm, and we found higher prevalence of adenovirus (PR 1.42, 95% CI 1.07, 1.87) in the nutrition-only arm (Table 9).

**Table 9:** Unadjusted prevalence ratios (PR) of detected bacteria, viruses, protozoa, and STHs, comparing intervention arms to control and single intervention arms to combined intervention.

|  | Compared to control arm |  |  | Compared to combined arm |  |
| --- | --- | --- | --- | --- | --- |
|  | Nutrition-only | Sanitation-only | Combined | Nutrition-only | Sanitation-only |
| **Any bacterium** | 1.06 (1.00, 1.11) | 0.99 (0.93, 1.05) | 1.03 (0.98, 1.08) | 1.03 (0.98, 1.08) | 0.96 (0.91, 1.02) |
| *Campylobacter* spp. | 1.08 (0.89, 1.32) | 1.09 (0.90, 1.32) | 1.18 (0.99, 1.40) | 0.92 (0.76, 1.12) | 0.93 (0.76, 1.12) |
| *C.diff* | 1.47 (0.95, 2.28) | 1.02 (0.63, 1.65) | 1.27 (0.84, 1.93) | 1.16 (0.75, 1.78) | 0.80 (0.50, 1.29) |
| EAEC | 1.08 (0.97, 1.20) | 1.00 (0.90, 1.12) | 1.04 (0.94, 1.14) | 1.04 (0.94, 1.16) | 0.97 (0.87, 1.08) |
| EPEC | 0.96 (0.85, 1.08) | 0.88 (0.78, 1.00) | 0.91 (0.81, 1.02) | 1.06 (0.93, 1.20) | 0.97 (0.85, 1.11) |
| aEPEC | 0.98 (0.84, 1.14) | 0.88 (0.75, 1.03) | 0.85 (0.73, 0.98) | 1.15 (0.97, 1.37) | 1.04 (0.88, 1.24) |
| tEPEC | 0.59 (0.34, 1.05) | 0.62 (0.36, 1.06) | 0.88 (0.57, 1.37) | 0.67 (0.37, 1.22) | 0.70 (0.39, 1.24) |
| ETEC | 1.07 (0.85, 1.35) | 0.90 (0.71, 1.15) | 0.99 (0.80, 1.22) | 1.08 (0.85, 1.38) | 0.91 (0.71, 1.17) |
| ETEC-LT | 1.18 (0.92, 1.53) | 0.98 (0.75, 1.29) | 0.95 (0.74, 1.21) | 1.25 (0.95, 1.65) | 1.04 (0.78, 1.38) |
| ETEC-ST | 1.26 (0.86, 1.86) | 1.10 (0.74, 1.63) | 1.42 (1.01, 2.00) | 0.89 (0.61, 1.29) | 0.77 (0.53, 1.13) |
| ETEC-LT/ST | 2.00 (1.19, 3.36) | 1.70 (1.00, 2.89) | 1.74 (1.06, 2.86) | 1.15 (0.71, 1.85) | 0.98 (0.60, 1.59) |
| *Salmonella* spp. | 1.04 (0.67, 1.62) | 0.65 (0.39, 1.08) | 1.01 (0.67, 1.51) | 1.03 (0.65, 1.64) | 0.64 (0.38, 1.09) |
| EIEC/*Shigella* spp. | 0.60 (0.39, 0.94) | 0.90 (0.62, 1.29) | 0.91 (0.65, 1.27) | 0.66 (0.42, 1.05) | 0.99 (0.67, 1.46) |
| STEC | 1.27 (0.79, 2.03) | 0.95 (0.57, 1.57) | 1.47 (0.97, 2.22) | 0.86 (0.55, 1.36) | 0.64 (0.39, 1.05) |
| **Any virus** | 1.09 (0.95, 1.26) | 1.03 (0.89, 1.20) | 1.16 (1.02, 1.31) | 0.94 (0.82, 1.09) | 0.89 (0.77, 1.03) |
| Adenovirus | 1.88 (1.41, 2.51) | 1.32 (0.96, 1.81) | 1.33 (0.99, 1.78) | 1.42 (1.07, 1.87) | 0.99 (0.73, 1.35) |
| Enterovirus | 0.93 (0.76, 1.13) | 0.92 (0.75, 1.12) | 1.12 (0.95, 1.32) | 0.82 (0.67, 1.01) | 0.82 (0.67, 1.00) |
| **Any protozoa** | 0.91 (0.68, 1.21) | 1.02 (0.78, 1.34) | 1.05 (0.82, 1.34) | 0.86 (0.64, 1.16) | 0.98 (0.74, 1.29) |
| *Giardia* | 0.91 (0.67, 1.23) | 1.09 (0.83, 1.43) | 1.02 (0.79, 1.33) | 0.89 (0.65, 1.22) | 1.06 (0.80, 1.42) |
| Gene targets with <5% prevalence were omitted from PR analyses: *V.cholera*, astrovirus, norovirus, rotavirus, sapovirus, *Cryptosporidium*, *Entamoeba*, and all STHs (*Ascaris*, *Trichuris*, *Ancylostoma*, and *Necator*). |  |  |  |  |  |

Generally, differences in mean gene quantities were consistent with prevalence differences (Table 9; Table 10). We detected lower concentrations of pathogen-associated genes in the nutrition-only and sanitation-only arms compared with the control arm; children in the nutrition-only arm carried lower quantities of STEC (−1.46 log_10_-copies, 95% CI -2.97, 0.06) and *Giardia* (−1.73 log_10_-copies, 95% CI -3.02, -0.44), and children in the sanitation-only arm carried lower quantities of EPEC (−0.54 log_10_-copies, 95% CI -1.17, 0.09) and STEC (−1.71 log_10_-copies, 95% CI -3.07, -0.34). There was no measurable difference in mean gene quantities between the combined and control arm. There was no significant difference in quantity of pathogen genes between treatment arms after adjusting for multiple comparisons^20^.

**Table 10:** Mean difference in log_10_-transformed gene copy estimates, comparing intervention arms to control and single intervention arms to combined intervention.

|  | Compared to control arm |  |  | Compared to combined arm |  |
| --- | --- | --- | --- | --- | --- |
|  | Nutrition-only | Sanitation-only | Combined | Nutrition-only | Sanitation-only |
| **Bacteria** |  |  |  |  |  |
| *CAMP* | 0.22 (-0.60 - 1.03) | -0.40 (-1.27 - 0.48) | -0.24 (-1.00 - 0.53) | 0.45 (-0.37 - 1.28) | -0.16 (-1.05 - 0.73) |
| *CDIF* | 0.31 (-1.14 - 1.75) | -0.37 (-1.92 - 1.19) | 0.19 (-1.16 - 1.54) | 0.11 (-1.29 - 1.52) | -0.56 (-2.08 - 0.96) |
| *EAEC aaic* | -0.38 (-1.35 - 0.60) | 0.35 (-0.52 - 1.22) | -0.28 (-1.08 - 0.52) | -0.10 (-1.11 - 0.92) | 0.63 (-0.28 - 1.54) |
| *EAEC aata* | -0.36 (-1.26 - 0.53) | -0.24 (-1.05 - 0.56) | 0.12 (-0.69 - 0.93) | -0.48 (-1.43 - 0.47) | -0.36 (-1.24 - 0.51) |
| *EPEC bfpA* | -1.08 (-3.47 - 1.31) | -1.13 (-3.16 - 0.89) | 1.15 (-0.38 - 2.67) | -2.23 (-4.57 - 0.11) | -2.28 (-4.24 - -0.32) |
| *EPEC eae* | -0.12 (-0.80 - 0.56) | -0.54 (-1.17 - 0.09) | 0.24 (-0.33 - 0.82) | -0.37 (-1.05 - 0.32) | -0.78 (-1.41 - -0.15) |
| *ETEC LT* | -0.62 (-1.64 - 0.40) | -0.47 (-1.56 - 0.61) | -0.03 (-1.06 - 1.00) | -0.59 (-1.65 - 0.47) | -0.44 (-1.57 - 0.68) |
| *ETEC stp* | 0.13 (-1.73 - 1.98) | 1.29 (-0.61 - 3.20) | 0.87 (-0.97 - 2.71) | -0.75 (-2.61 - 1.11) | 0.42 (-1.49 - 2.33) |
| *SALM* | 1.42 (-0.04 - 2.87) | 0.27 (-0.97 - 1.52) | 0.80 (-0.33 - 1.93) | 0.62 (-0.93 - 2.17) | -0.53 (-1.89 - 0.83) |
| *IPAH* | 0.22 (-1.33 - 1.77) | -0.28 (-1.68 - 1.13) | 1.17 (0.07 - 2.26) | -0.95 (-2.43 - 0.53) | -1.44 (-2.77 - -0.11) |
| *STEC1* | -1.46 (-2.97 - 0.06) | -1.71 (-3.07 - -0.34) | -0.00 (-1.49 - 1.49) | -1.46 (-2.84 - -0.07) | -1.70 (-2.92 - -0.49) |
| *STEC2* | -0.20 (-1.45 - 1.04) | 0.09 (-1.20 - 1.37) | 0.72 (-0.47 - 1.91) | -0.92 (-2.06 - 0.22) | -0.63 (-1.82 - 0.56) |
| **Viruses** |  |  |  |  |  |
| *ADEV* | 0.50 (-0.48 - 1.49) | 0.67 (-0.32 - 1.67) | 0.48 (-0.42 - 1.38) | 0.03 (-0.96 - 1.02) | 0.20 (-0.80 - 1.20) |
| *ENTV* | -0.40 (-1.09 - 0.28) | -0.35 (-0.97 - 0.26) | -0.26 (-0.77 - 0.24) | -0.14 (-0.84 - 0.56) | -0.09 (-0.72 - 0.54) |
| **Protozoa** |  |  |  |  |  |
| *GIAR* | -1.73 (-3.02 - -0.44) | 0.23 (-1.16 - 1.62) | -0.14 (-1.47 - 1.18) | -1.58 (-3.06 - -0.11) | 0.37 (-1.19 - 1.93) |
| Gene targets with <5% prevalence were omitted from PR analyses: *V.cholera*, astrovirus, norovirus, rotavirus, sapovirus, *Cryptosporidium*, *Entamoeba*, and all STHs (*Ascaris*, *Trichuris*, *Ancylostoma*, and *Necator*). |  |  |  |  |  |

## Discussion

We found a modest effect on growth from the nutrition-only intervention and a greater effect in the combined intervention arm that was likely attributable to the nutrition intervention alone. By contrast, the sanitation intervention alone was not associated with growth improvements, relative to control conditions, and demonstrated significantly poorer growth than the combined intervention arm. The similar impacts on linear growth between the nutrition-only and combined intervention arms were consistent with the observed linear growth improvements being attributable primarily to the nutrition intervention alone, further suggesting that the addition of this sanitation intervention to the nutrition intervention did not produce synergistic effects. Intermediate outcomes of meal frequency and dietary diversity were similar between arms, so the observed effects may have been attributable to other elements of the nutrition or combined intervention not captured by these measures. We observed no meaningful differences between arms with respect to secondary outcome measures of WAZ, WLZ, stunting, wasting, underweight status, diarrhoea, mortality, pathogen prevalence, pathogen co-detection rate, or pathogen gene copy quantity. Although molecular detection of a specific pathogen in stool does not necessarily signal active enteric infection, potential for disease, or direct effects on the individual, it does unambiguously indicate prior exposure to that pathogen. Our specific pathogen targets were selected *a priori* based on a range of globally observed enteric pathogens and may not fully capture the relevant enteric pathogens in rural Cambodia; however, the consistently high pathogen prevalence across all treatment arms suggests the suite of interventions assessed in this trial did not prevent environmental exposure to enteric pathogens.

Our findings are consistent with results from several recent randomised factorial WASH and nutrition efficacy trials reporting protective effects of combined/integrated interventions and null effects of WASH alone on child growth outcomes^3,4,9^. A small number of experimental trials^24^ and many observational studies^8,10^ have reported increases in child growth and reductions in stunting prevalence with improvements in sanitation coverage and commensurate reductions in OD; among the latter, unmeasured confounding is a likely explanation for observed effects^24^.

While gains in the proportion of the population self-reporting access to sanitation were highest in the sanitation only arm (+25 pp), these were only modestly higher than the gains for the control (+19 pp) and nutrition arms (+14 pp); furthermore, sanitation coverage gains for the combined intervention matched the control arm at +19 pp. Comparable secular trends of increasing sanitation coverage in Cambodia have been documented previously: the percentage of children younger than five years of age with access to an improved sanitation facility increased from 5% in 2000 to 17% in 2005, 29% in 2010, and 54% in 2014, the most recent nationally representative DHS survey^8,23^. Correspondingly swift improvements have been documented specifically in rural areas, where access to any sanitation facility increased from 30% in 2010 to 44% in 2014 and improved sanitation coverage rose from 27% to 43%^8,23^. The rapid pace of WASH development in rural Cambodia makes it challenging to measure the impact of specific programs. However, the lack of differences in other sanitation intervention adherence indicators suggests low overall uptake of the sanitation intervention and only modest increases in sanitation coverage attributable to the intervention, which were likely insufficient to reduce community exposure and transmission. Zoonotic transmission from domestic animals, for instance, was not addressed by this or many other WASH trials, as indicated by nearly 80% of households across all treatment arms lacking access to an area free of animals for children to safely play^25^.

The frequency and intensity of contact from program promoters was much greater in the nutrition intervention than the sanitation intervention. Recipients of the nutrition intervention participated in monthly activities, whereas the sanitation intervention consisted of one triggering session with infrequent follow-up visits. The lower contact frequency may explain the discrepancy in intervention adherence. Both arms receiving the nutrition programming reported higher levels of participation in the key intervention activities—including sanitation intervention activities—suggesting higher adherence to the nutrition intervention than the sanitation intervention. Self-reported CLTS participation rates were equally low in both the sanitation-only and control arms at 6%, while 14% of nutrition-only recipients and 25% of combined intervention recipients reported CLTS participation. The comparatively elevated CLTS participation in the nutrition-only arm may reflect biases embedded in the self-reporting process; given the 28+ months that had elapsed since the initial CLTS triggering session and the infrequency of CLTS follow-up visits, households that only received the sanitation intervention may have been less likely to recall programming of any kind than households that participated in the more frequent and intense nutrition intervention activities. Furthermore, the “Growth Together” SBCC campaign, which promoted 13 core health, nutrition, sanitation, and hygiene practices, was fully incorporated into all intervention activities across the three intervention arms, meaning households receiving the higher intensity nutrition programming also encountered the associated SBCC sanitation messaging much more frequently than households in the sanitation-only arm. The SBCC campaign was also promoted nationally on television, such that households in the control arms may have been nearly as exposed to its content as sanitation-only households, while the nutrition-only and combined intervention arms received substantial in-person promotion of the SBCC campaign messaging.

Due to the nature of the interventions and resource considerations, all trial outcomes were assessed during a single survey round conducted 28 months after initiating the intervention programming, which introduced some limitations. Growth and pathogen outcomes were assessed in children from one to 28 months of age, meaning that older children received the treatments for a longer duration but were initially exposed to less mature intervention conditions than the younger children born later. The timing of outcome ascertainment also precluded detecting effects that may only be realized later in childhood, such as potentially rapid catch-up growth after 24 months of age that may reverse earlier growth faltering^26^. While a focus on the first 1,000 days is justified^2^, investigation of growth and growth-promoting factors after this window may provide additional insight on improved WASH practices and their role in supporting long-term development and health.

While molecular detection of a specific pathogen in stool unambiguously indicates prior exposure to that pathogen, our data do not necessarily indicate active enteric infection, potential for disease, or direct effects on the individual. We were limited by the suite of pathogen targets selected on our custom TAC assay; we selected these targets *a priori* based on a range of globally observed enteric pathogens, but we cannot know whether these were the most important enteric pathogens in rural Cambodia. There is also evidence that the *invA* gene, which was selected for *Salmonella* spp. detection, is not specific to *Salmonella enterica* and suggest the consideration of other genes, such as *ttrA/C*, for reliable detection of *S. enterica*^27^. It is highly plausible that this sanitation intervention simply failed to sufficiently reduce environmental exposure to enteric pathogens. For example, only 22% of households in our survey were observed to have a child play environment free of animals, with little difference between treatment and control arms; this is a transmission pathway that our trial and many other WASH trials have not addressed^25^. The trial design is predicated on the theory that gains in sanitation coverage may lead to improved growth outcomes in children via reductions in the transmission of enteric infection and disease, though links between sanitation coverage and specific outcomes are poorly understood in high-burden settings. The change in community coverage in this trial was limited and likely insufficient to reduce community exposure and transmission.

There are a few key observations from this study that should be considered in future interventions and effectiveness trials of comparable interventions. Increased frequency, duration, and intensity of CLTS programming could have resulted in greater uptake of sanitation in target communes. Despite the sanitation coverage gains observed in the sanitation-only arm, much of which may have been as a result of the sanitation intervention, we are unable to attribute beneficial effects—i.e., measurable differences in prespecified outcomes—to the sanitation intervention due to the high sanitation gains also observed in the control arm. There may have been other benefits of sanitation gains that were not measured, including in safety and broader measures of well-being^28^. Future trials may also include additional objective outcome measures, including intermediate measures of environmental contamination that are on the causal pathway between interventions and exposures.

Our work is consistent with a growing body of research reporting high prevalence of enteric pathogen exposure in early childhood, which may lead to long-term effects on health^15,18,29,30^. Reducing these exposures in high-burden settings requires transformative interventions that have the potential to dramatically reduce direct and indirect contact with all faeces, including animal faeces^24^, across multiple pathways.

## Data Availability

The data that support the findings of this study are openly available in OSF at DOI 10.17605/OSF.IO/PNU4A.

https://www.doi.org/10.17605/OSF.IO/PNU4A

## Other Information

### Trial Registry

The trial is registered with ISRCTN Registry (<u>ISRCTN77820875</u>).

### Protocol

The National Ethics Committee for Health Research in the Cambodian Ministry of Health reviewed and approved the protocols (NECHR #110) prior to the start of data collection. The study also received approvals from the Institutional Review Board at Georgia Institute of Technology (Ref: H19286) and from New England IRB (IRB#: 120190186).

## Acknowledgements

We thank the staff and participants of the study for their important contributions. We also acknowledge contributions by Caroline Akerley, Farran Bush, Isabelle du Plessis, and Juliann Pham.

## Author contributions

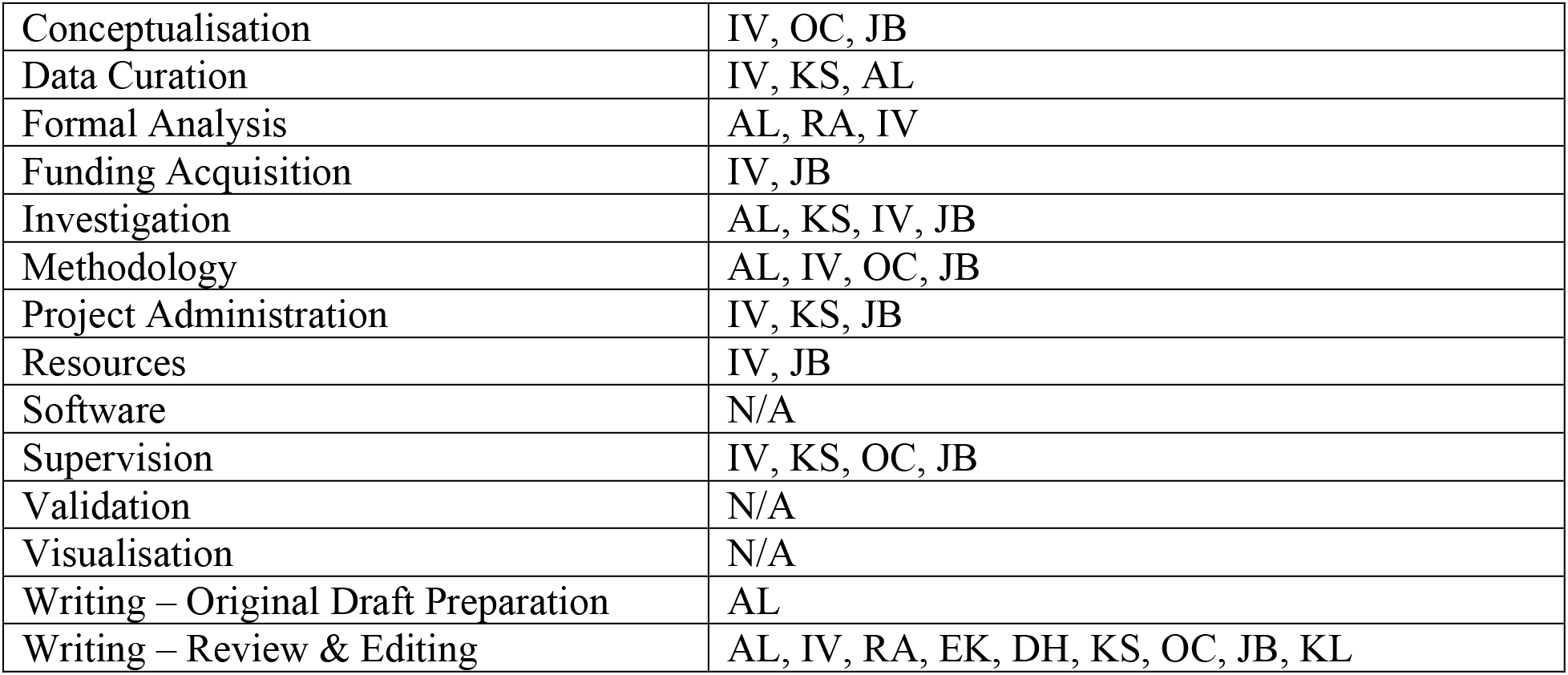

