## Supplementary Materials for "Independent and combined effects of nutrition and sanitation interventions on enteric pathogen carriage and child growth in rural Cambodia: a factorial cluster-randomised controlled trial"

**Table of Contents**

**S1: Intervention procedures** .......... **2**
**S2: Data collection** .......... **6**
**S3: Sample size and power calculations** .......... **8**
**S4: Anthropometry protocols** .......... **10**
**S5: Nucleic acid extraction and PCR procedures** .......... **11**
**S6: TaqMan Array Card performance and standard curve parameters** .......... **12**
**S7: List of primers and probes in custom-TAC.** .......... **13**
**S8: qPCR assay validation** .......... **15**
**S9: Process evaluation indicators** .......... **16**
**S10: Intervention adherence** .......... **17**
**S11: Adjusted analyses** .......... **18**
**S12: Age-stratified pathogen prevalence** .......... **22**
**S13: Pre-intervention sanitation coverage** .......... **25**
**S14: CONSORT checklist for RCTs** .......... **27**
**Additional trial limitations [probably delete this]** ..........Error! Bookmark not defined.
**References** .......... **30**

**S1: Intervention procedures**

Nutrition Interventions

The nutrition interventions consisted primarily of complementary feeding activities and education through community-based delivery platforms, and conditional cash transfers, vouchers, and social and behavioural change (SBCC) linked to the adoption and utilization of key health and nutrition practices, services, and products.

The Community Nutrition component used evidence-based integrated nutrition interventions for the "first 1,000 days" of life. Village Health Support Groups (VHSGs), supervised by health workers and Commune Councils for Women and Children (CCWC), sought to improve childcare and development at multiple levels: individual, family, and community. Five core activities comprised the community initiative designed to prevent malnutrition:

1) Community Dialogues: This activity was led quarterly by the Village Chief and VHSGs. The community gathered to talk, decide, and take action together to support all children to grow healthy. Communities reviewed progress of creating a healthy environment, discussed one key action to jointly address challenges, and decided together how everyone can come together to achieve this action.
2) Caregiver Group Education Sessions: Caregiver Groups were peer-led groups of women who use a 13- session experiential learning manual following each of the key behaviours promoted by the program implementers. Program staff trained two members per group to facilitate monthly sessions for their group, with support from elder women in the community and trained Community Agents.
3) Growth Monitoring and Promotion: VHSGs monitored every child every month. Children who were sick or not growing well were referred to health centres or referral hospitals, as appropriate, and followed up at home after treatment.
4) Home visits: VHSGs and Mother Support Group (MSG) members provided tailored interpersonal communication during home visits to promote childcare and feeding practices, home hygiene, and proper use of latrines and handwashing stations. Home visits were conducted for pregnant women, caregivers of children 9-11 months old and caregivers of children not growing well.
5) Village Fairs: This activity was held twice a year for each village. Village fairs offered women and their families' hands-on learning experiences that bring together health/nutrition, WASH and agriculture using games, demonstrations and practice, interactive discussions and latrine marketing and sales by local participating sanitation suppliers.

CCT acted as a social safety net mechanism for poor "first 1,000 days" families, serving as an incentive for women to access services, practice specific behaviours, and overcome constraints related to poverty. Eligible families (based on poverty status) could receive up to six payments, for a total of $65 over the first 1,000 days of a child's life, which were transferred directly into women's bank accounts after completed use of health and nutrition services.

1) First transfer: $12.50 at 1 month postpartum. Conditions: At least four antenatal care visits, delivery in a health centre, and at least two postnatal care visits.
2) Remaining five transfers: $10 for the second to fifth transfers and $12.50 for the last transfer over the next 23 months postpartum. Conditions: Monthly monitoring of

children's growth through Growth Monitoring and Promotion (GMP) at the health centres or in the community, and handwashing station at home.

Vouchers served as another mechanism to encourage demand and overcome access constraints related to poverty. Vouchers were distributed to poor "first 1,000 days" families in communes where the CCT is implemented and is redeemable for discounts on water filters ($5 co-payment) and two food baskets ($5 co-payment). Vouchers were only distributed in combined intervention groups.

SBCC consisted of media and materials to promote key behaviours in health/nutrition, sanitation/hygiene, and agriculture. The project's SBCC framework was grounded in evidence of what works in social and behaviour change and foundational work done by program implementers the year before the start of the study. On the nutrition side, SBCC supported all the Community Nutrition activities, described above, and was implemented by community change agents (VHSGs and caregiver peer groups).

1) Grow Together: The campaign focused on 13 key stunting prevention behaviours (Figure below) spanning health, nutrition, WASH, and agriculture to stimulate relevant actions for children to grow and reach their full potential. It was not possible to exclude the WASH messaging from the Grow Together campaign, so caregivers received information on all 13 behaviours as part of the nutrition programming.

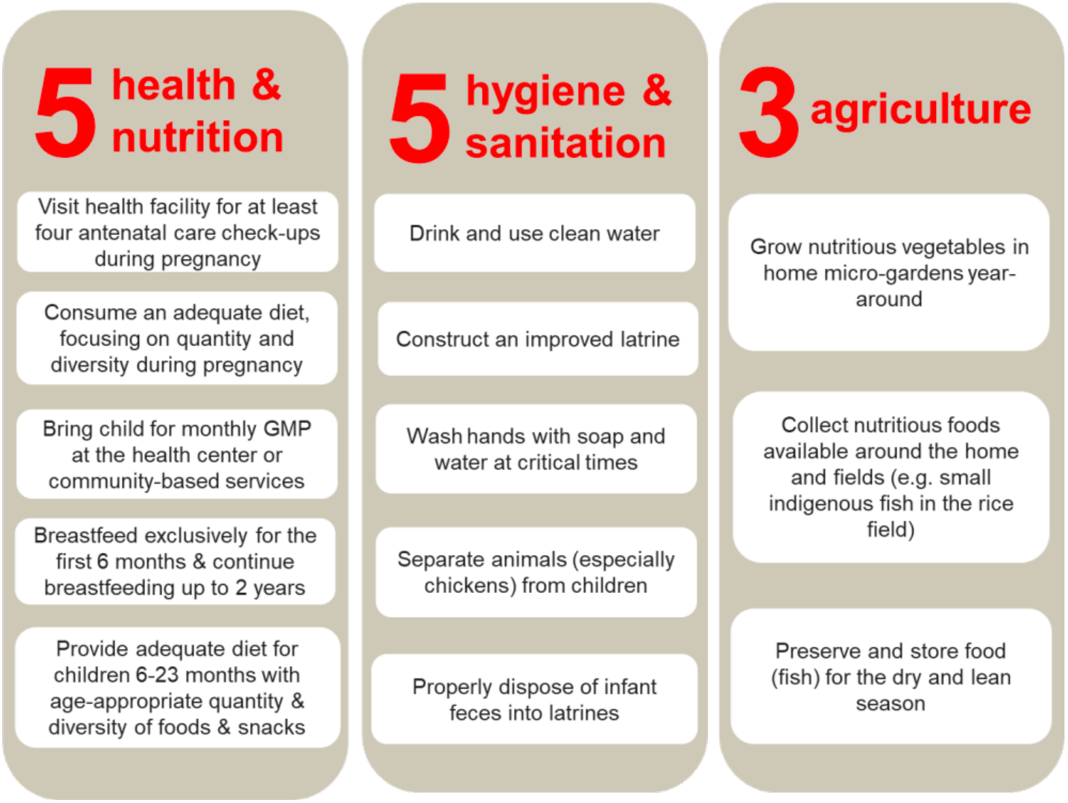


2) To complement the print materials, the SBCC media plan included three television spots including the foundational Grow Together TV spot, latrine construction and Small Fish Powder; 13 "soundbites"; an advocacy package for local leaders; and more than 20 print materials carrying the same "look and feel" to link to core values and motivations to take action.

3) The “first 1,000 days” family SBCC package was centered on a Family Commitment Card enumerating the critical practices and allowing families to prioritize behaviours and visualize successes and gaps. As the Family Card filled with accomplishments, the family was recognized as a growth champion with a child book and other incentives to mark its accomplishment. A behaviour wheel checklist to guide home visits showing health/nutrition and sanitation/hygiene practices supplemented the Family Card.

Sanitation Interventions

The sanitation interventions consisted primarily of community-led total sanitation (CLTS), coupled with supply-side support for sanitation and hygiene products, latrine vouchers, and SBCC on hygiene practices.

CLTS aimed to achieve sustained behaviour change through the process of community “triggering” leading to spontaneous and long-term abandonment of open defecation practices. This one-time triggering event was conducted in collaboration with the Ministry of Rural Development and provincial and district departments of rural development. In alignment with national Open Defecation Free certification guidelines, CLTS covered entire villages to minimize the risk of fecal-oral contamination for all children. Following CLTS triggering, program staff monitored the commitments of families and communities through door-to-door visits. These visits took place at least 5 times per village and were used to also raise awareness and create demand for sanitation/latrines.

Latrine vouchers were a targeted subsidy to poor households in villages that reached 75% sanitation coverage. Vouchers were redeemable for a discount on latrine materials ($15 co-payment). In the combined-intervention villages, latrine vouchers were initially linked to the CCT program, and so were only offered to those beneficiaries. However, this requirement was phased out and latrine vouchers were eventually available to all poor households in eligible villages.

Supply-side support consisted of collaborating with private and public sector actors to develop locally sensitive market-oriented approaches for the integrated business service centres around “first 1,000 days” products and services. Program staff encouraged knowledge sharing across small- and medium-sized enterprises (SMEs) as well as utilized existing resource centres and agencies to develop the capacity of SMEs for effective service delivery and to increase their outreach to poor and relatively remote areas. Program staff identified a number of successful businesses within or outside the project area and organized interfaces between new and existing businesses to give mutual learning opportunities between SMEs and develop linkages for possible collaboration. In addition, suppliers were linked to communes where CLTS triggering had occurred so they could follow-up with households that committed to purchasing or building latrines.

SBCC on the sanitation side consisted of sanitation campaigns in primary schools to ensure children become agents of change and carry new behaviours home. As change agents, children have the potential to convince their families to construct latrines or purchase a handwashing station, and to use them.

Additional notes on intervention procedures

There were differences in frequency and intensity of contact from program promoters across intervention arms, possibly resulting in reduced impact of the relatively light-touch sanitation intervention. Arms receiving nutrition intervention participated in monthly activities, whereas arms receiving sanitation intervention participated in one triggering session with few and infrequent follow-up visits. Thirteen core health/nutrition and sanitation/hygiene behaviours were promoted as part as the “Growth Together” SBCC campaign. The campaign was broadcasted on television indiscriminately across the country – and subsequently across treatment and control arms. The core behaviours were also promoted during all intervention activities, resulting in higher intensity of programming (including promotion of sanitation and hygiene practices) in the nutrition arms. The lower frequency in contact may explain the discrepancy in intervention adherence. The nutrition arms reported higher levels of participation in the key intervention activities, suggesting higher adherence of the nutrition intervention compared to the sanitation intervention. Similarly, those in the sanitation-only arm reported CLTS participation rates no different from the control arm (6% and 6%), compared with self-reported CLTS participation in the nutrition and combined arms (14% and 25%; **Error! Reference source not found.**; **Error! Reference source not found.**). The high self-reported CLTS participation in the nutrition-only arm compared to the control arm may reflect biases embedded in the self-reporting process, especially when considering the time elapsed since the initial CLTS interventions took place (28+ months prior) and how infrequently CLTS contact occurred relative to nutrition intervention. Households that already had access to sanitation may not have engaged with the CLTS programming, the survey respondent may not have been aware of or may not recall specific activities, or other reporting biases could have played a role. The greater frequency and intensity of contact between the interventions and the respondents in the nutrition arms may have resulted in greater apparent recall of programming of any kind in this arm, possibly increasing reporting and observer biases; participants were not masked to intervention status due to the nature of the interventions. It is also possible that there were other nutrition- and/or WASH-related outreach efforts from actors external to the intervention program. We included observable indicators in addition to self-reported measures as indicators of intervention adherence, which included direct observations of sanitation facilities and domestic hygiene status (e.g., faeces in the play environment of children).

**S2: Data collection**
A primary survey, based mainly on validated Cambodia DHS questionnaires and piloted in adjacent districts to the study area, was conducted to assess household and child-level risk factors of children under 28 months of age.

Enumerators completed in-home interviews, in the Khmer language, with the primary caregiver of children between 1 to 28 months of age in the household. Field staff asked caregivers questions about basic household member information; breastfeeding, health, and diet of the target children; hygiene, water and sanitation practices; pregnancies and child births of the caregiver; intervention exposure and participation; household WASH conditions; and household assets/characteristics to construct wealth scores. Child height and weight were measured by trained paired enumerators following guidelines from the Food and Nutrition Technical Assistance project (FANTA)[1].

For nutrition-related data collection, we included the infant and young children feeding indicators suggested by the World Health Organisation (WHO), which include minimum dietary diversity, minimum meal frequency, and minimum adequate diet[2]. WHO dietary diversity score consists of categorising solid foods into seven food groups, including: grains, legumes/nuts, dairy, flesh meat, eggs, vitamin-A-rich fruits and vegetables, and other fruits and vegetables. To suit the Cambodian context, the evaluation team asked additional questions on the types of fish and other wild animals consumed, which are included in the flesh meat group. The dietary diversity score is on a scale from 0 – 7 and determined based on the number of food groups the caregiver reported to have fed the child in the last 24 hours; minimum dietary diversity is defined as having received food from four or more food groups (or a dietary diversity score greater than or equal to four). Minimum meal frequency is defined by the frequency of solid and semi-solid foods received based on a child's age and whether the child is breastfed. The minimum number of times breastfed children should receive solid, semi-solid, or soft foods varies with age (2 times if 6–8 months and 3 times if 9–23 months). The minimum number of times non-breastfed children should receive solid, semi-solid, or soft foods, including milk, is 4 times for all children 6–23 months.

For sanitation-related data collection, we included household WASH indicators and environmental hygiene indicators. Household WASH indicators included drinking water source, access, and treatment; handwashing station access; sanitation facilities access; and disposal of child stools. Environmental hygiene indicators included presence of human stools, animal faeces, animals, and garbage in child's play environment.

As part of a supplemental analysis to assess the effects of key community-level WASH indicators (sanitation coverage and rates of open defecation), a secondary survey was conducted in households randomly selected in the same areas (three households per village) and irrespective of whether there was a child living in the household. Given the oversampling of households with children under 28 months of age, post-stratification weights were used to get a representative sample of the population. Sampling weights were calculated as follows: first, we estimated the proportion of households with children under 28 months of age at the village-level by creating a list of eligible children with the village chief and VHSG. This estimate was then divided by the proportion of sampled households with children under 28 months of age at each village to yield the sampling weight for each household from the main sample. For the three additional households, the sampling weights were calculated by dividing the remaining proportion of total households at the village level by the proportion of sampled households at each village. This resulted in underweighting the households with children under 28 months of age and overweighting the supplemental households.

**S3: Sample size and power calculations**
Sample size was chosen to balance the size of the study and the minimum detectable difference between arms. Increased allocation of eligible communes to the control arm was stipulated to enhance statistical efficiency of multiple hypothesis testing, resulting in 19 communes assigned to the control arm (one-third of the total) and 13 to each intervention arm[3]. Power calculations used α=0.05, power=0.8, mean LAZ estimate (prior to intervention rollout) of -0.96 with a standard deviation of 1.19, intra-cluster correlation of 0.014 on the LAZ outcome measure at the commune level, and a two-sided test for a two-sample comparison of means. LAZ calculations used a standard equation assuming a single, post-treatment measurement at 2 years, resulting in a total of 4,015 households consisting of 73 observations per commune. These sample size calculations suggest that this study had sufficient power to detect a minimum detectable effect size (MDES) of 0.19 for differences in the LAZ scores between treatment arms and a MDES of 0.18 for differences between each treatment arm and the control arm, similar to other trials[4]. An MDES of 0.19 translates to a 23.4% change in LAZ score between treatment arms; an MDES of 0.18 translates to a 22.2% change in LAZ scores between treatment and control arms[5]. While empirical evidence to serve as an adequate basis for the MDES was limited, another large factorial WASH and nutrition trial targeted a similar LAZ MDES of 0.18 between treatment arms and a MDES of 0.15 in mean LAZ scores between treatment and control arms[6,7].

The following sample size calculations for the primary outcome, difference in mean HAZ scores between treatment groups, were conducted based on different ICC scenarios. This is revised to account for a drop of three treatment communes after randomization occurred. Power calculations assume α=0.05, power=0.8, mean baseline HAZ estimate of -1.637 with a standard deviation of 1.286, and a two-sided test for a two-sample comparison of means. HAZ calculations use a standard equation assuming a single, post-treatment measurement at 2 years.

| MDES between treatment groups | Subjects per cluster (commune) | Estimated total number of subjects required (all 4 arms) |
|---|---|---|
| | ICC=0.01 | |
| 0.15 | 155 | 8,525 |
| 0.16 | 115 | 6,325 |
| 0.17 | 90 | 4,950 |
| **0.18** | **73** | **4,015** |
| 0.19 | 61 | 3,355 |
| 0.2 | 52 | 2,860 |
| | ICC=0.015 | |
| 0.15 | 690 | 37,950 |
| 0.16 | 268 | 14,740 |
| 0.17 | 162 | 8,910 |
| 0.18 | 114 | 6,270 |
| 0.19 | 87 | 4,785 |
| 0.2 | 70 | 3,850 |
| | ICC=0.02 | |

| | | |
|---|---|---|
| 0.15 | NA | NA |
| 0.16 | NA | NA |
| 0.17 | 876 | 48,180 |
| 0.18 | 268 | 14,740 |
| 0.19 | 155 | 8,525 |
| 0.2 | 107 | 5,885 |

For the supplemental analysis on community-level WASH indicators, the required sample size was calculated based on a conventional approach for proportions to collect reliable point-estimates of sanitation coverage at the group level, at the 95 percent confidence level with a margin of error of +/-5 percent.

$$N = p(1-p) * \left[\frac{Z}{ME}\right]^2 * 4 = 0.408(1-0.592) * \left[\frac{1.96}{0.05}\right]^2 * 4 = 1{,}024$$

where:
$p$ = proportion of sanitation coverage of 0.408, estimated using DHS 2014 data
$Z$ = 1.96 (for 95% confidence level)
$ME$ = margin of error of +/-5%

Given the 491 villages, sample size was rounded up to three additional randomly selected households per village, for a target total of 1,473 additional households, as shown in the table below.

Required Sample Size

| Provinces | Communes | Villages | HHs Main Sample | HHs Secondary Sample |
|---|---|---|---|---|
| Battambang | 22 | 180 | 1,606 | 540 |
| Pursat | 6 | 83 | 438 | 249 |
| Siem Reap | 27 | 228 | 1,971 | 684 |
| **Total** | **55** | **491** | **4,015** | **1,473** |

**S4: Anthropometry protocols**

Anthropometric measurement is comprised of weight and length. Weight was measured using Uniscale (UNICEF recommended scale) in Kilogram with precision to one decimal point. Length was measured using a length board (UNICEF / WFP recommended) in Centimetre with precision to one decimal point. Two data measurements were required, one from the measurement taker and another one from an assistant. The measurement procedure followed FANTA Guidelines:

- Weight measurement:
    - *Preparation*: Ensure enough material is available for measurement (scale, battery, pen, tissue, record form, and age calculation form) with proper function. Ensure that the scale is positioned in a plate and smooth surface. Measurement taker is on the right hand of mother/caregiver while assistant is in front of mother/caregiver. Ensure that children dress light clothes with no cap or shoes. Assistant helps mother/caregiver in carrying the child and asks mother/caregiver to go on to the scale after proper functioning.
    - *During Measurement*: Request mother/caregiver to stand on the scale, inform the measurement result loudly, press button to measure child, hand the child to mother/caregiver after scale functioning, read weight of child out loud so that assistant can record the measurement.
    - *Second Measurement*: Request mother/caregiver to step off the scale. Repeat the measurement steps. Record second measurement.

- Length measurement:
    - *Preparation*: Prepare length board on a plate and smooth surface. Ensure length board stability, take off shoe and cap from child. Check measurement level on the length board, and ensure the record form.
    - *During Measurement*: Lay child on his back on the board, check head, eye, shoulder, hand, buttock, knees and heel. Make sure body is in proper position and still. Measurement must be read to the nearest of 0.1 cm. Repeat the measurement one more time to ensure accuracy of reading. If the two measurements are different by more than 1.0 cm, then a third measurement is taken.

- Following the weighing and length measurements, any child who is classified as severely malnourished is referred to the health clinic.

- Training: The enumerators were trained on the protocols to follow and how to calibrate equipment. Tested on accurate recording of length measurements. Hands-on practice in pairs and then we did a standardization exercise where the entire team is tested on their ability to measure child length accurately and precisely. Measurers had to meet the accuracy and precision threshold to pass and be hired as enumerators for data collection.

- Field supervision: The anthropometry specialist was present in the field during the entire baseline phase, accompanying enumerators to ensure proper technique with the height and weight measurements and recording.

**S5: Nucleic acid extraction and PCR procedures**

Stool samples were collected and preserved in duplicate using Zymo DNA/RNA Shield buffer (Zymo Research, Irvine, CA) at 1:1 by volume and stored in -20°C until extraction. A subset of stool samples were randomly selected for extraction and molecular analysis. Our extraction protocol was adapted from the xTAG Gastrointestinal Pathogen Panel (GPP; Luminex Molecular Diagnostics, Toronto, ON, Canada) protocol for pre-treatment and the QIAamp 96 Virus QIAcube HT (Qiagen, Germany) protocol for remaining extraction procedures[8]. Briefly, 200 mg solid (or 200 uL if liquid) preserved stool was combined with 1000 uL of Buffer ASL (Qiagen, Germany) in an SK38 soil grinding tube (Bertin, Rockville, MD), vortexed for 5 minutes (Vortex Genie 2, Scientific Industries, Bohemia, NY), incubated at room temperature for 10 minutes, and centrifuged at 12,000g for 2 minutes (Eppendorf, Enfield, CT). 200 uL of supernatant was used for total nucleic acid extraction following the QIAamp 96 Virus QIAcube HT protocol. We assayed total nucleic acids using a custom-developed TaqMan Array Card (TAC; ThermoFisher Scientific, Waltham, VA) – a compartmentalised probe-based quantitative real-time polymerase chain reaction (PCR) assay for 30 enteric pathogen genes using individual assays validated in previously published literature in TAC format[9,10]. PCR cycling conditions were also adapted from previous work[9,10]. Details on specific targets, assays, assay validation, and other analytical metadata are included in Supporting Information.

We collected 4,114 stools in totla and assessed enteric pathogen-associated gene targets in 1,745 for molecular analysis using multiple-target PCR for presence of gene targets associated with key enteric pathogens (bacteria, viruses, protozoa, and STH). We omitted 125 samples due to lack of amplification of one or more of three controls (phHPV as DNA control; MS2 as RNA control; manufacturer internal positive control) or due to unstable noise in amplification curves. 1,620 samples were included in the final dataset.

**S6: TaqMan Array Card performance and standard curve parameters**

| Target | Target gene | Slope | Y-intercept | R2 | Efficiency | LLOD (GC/rxn)* |
|---|---|---|---|---|---|---|
| Enteric bacterial 16S | 16S | -3.613 | 42.29 | 0.960 | 89% | 10 |
| pan-Adenovirus | hexon | -3.372 | 38.58 | 0.994 | 98% | 10 |
| *Ancylostoma duodenale* | *ITS2* | -3.506 | 41.68 | 0.994 | 93% | 10 |
| *Ascaris lumbricoides* | *ITS1* | -3.479 | 40.72 | 0.992 | 94% | 10 |
| Astrovirus | capsid | -3.337 | 37.89 | 0.997 | 99% | 10 |
| *Campylobacter jejuni* | cadF | -3.562 | 40.22 | 0.999 | 91% | 10 |
| *Clostridium difficile* | tcdB | -3.427 | 38.25 | 1.000 | 96% | 10 |
| *Cryptosporidium parvum* | *LIB13* | -3.505 | 40.17 | 0.999 | 93% | 10 |
| *Cryptosporidium hominis* | *LIB13* | -3.433 | 39.49 | 0.999 | 96% | 10 |
| EAEC (aaic) | *aaic* | -3.342 | 36.18 | 0.997 | 99% | 1 |
| EAEC (aata) | *aata* | -3.221 | 35.10 | 0.987 | 104% | 1 |
| *Entamoeba histolytica* | 18S rRNA | -3.406 | 40.42 | 0.993 | 97% | 10 |
| Enterovirus | 5'UTR | -3.396 | 38.94 | 0.999 | 97% | 10 |
| EPEC (bfpa) | *bfpa* | -3.380 | 37.81 | 0.994 | 98% | 10 |
| EPEC (eae) | *eae* | -3.391 | 38.20 | 0.995 | 97% | 10 |
| ETEC (LT) | LT | -3.591 | 39.38 | 0.988 | 90% | 1 |
| ETEC (STh) | STh | -3.428 | 38.46 | 0.996 | 96% | 10 |
| ETEC (STp) | STp | -3.377 | 37.75 | 0.994 | 98% | 10 |
| *Giardia* spp. | 18S rRNA | -3.412 | 40.17 | 0.999 | 96% | 10 |
| EIEC/*Shigella* spp. | *ipaH* | -3.332 | 38.14 | 0.999 | 100% | 10 |
| *Necator americanus* | *ITS2* | -3.434 | 40.55 | 0.994 | 96% | 10 |
| Norovirus GI | ORF1-2 | -3.457 | 39.85 | 1.000 | 95% | 10 |
| Norovirus GII | ORF1-2 | -3.387 | 38.53 | 0.999 | 97% | 10 |
| Rotavirus | NSP3 | -3.624 | 41.79 | 0.998 | 89% | 10 |
| *Salmonella* spp. | *invA* | -3.446 | 40.67 | 0.996 | 95% | 10 |
| Sapovirus I | RdRp | -3.392 | 39.22 | 0.998 | 97% | 1 |
| Sapovirus IV | RdRp | -3.384 | 39.00 | 0.998 | 97% | 10 |
| STEC | *stx1* | -3.397 | 38.49 | 0.998 | 97% | 1 |
| STEC | *stx2* | -3.396 | 38.53 | 0.997 | 97% | 1 |
| *Trichuris trichiura* | 18S rRNA | -3.307 | 40.31 | 0.999 | 101% | 100 |
| *Vibrio cholerae* | *hlyA* | -3.418 | 41.00 | 0.999 | 96% | 100 |

*Lower limit of detection estimated by assuming Cq cutoff of 35[11]

**S7: List of primers and probes in custom-TAC.**
All sequences were based on cited references.

| | Organism | Target gene | Forward Sequence #1 (5'-3') | Reverse Sequence #1 (5'-3') | Probe Sequence #1 (5'-3') | References |
|---|---|---|---|---|---|---|
| Virus | Astrovirus | capsid | CAGTTGCTTGCTGCGTTCA | CTTGCTAGCCATCACACTTCT | CACAGAAGAGCAACTCCATCGC | [1] |
| | Enterovirus | 5'UTR | CCCTGAATGCGGCTAATCC | GCGATTGTCACCATWAGCAG | CCGACTACTTTGGGWGTCCGT | [2] |
| | Norovirus GI | ORF1-2 | CGYTGGATGCGNTTYCATGA | CTTAGACGCCATCATCATTYAC | TGGACAGGAGATCGC | [2] |
| | Norovirus GII | ORF1-2 | CARGARBCNATGTTYAGRTGGATGAG | TCGACGCCATCTTCATTCACA | TGGGAGGGCGATCGCAATCT | [1] |
| | Sapovirus | RdRp | GAYCAGGCTCTCGCYACCTAC | CCCTCCATYTCAAACACTA | CYTGGTTCATAGGTGGTRCAG | [1] |
| | | RdRp | TTTGAACAAGCTGTGGCATGCTAC | CCCTCCATYTCAAACACTA | CAGCTGGTACATTGGTGGCAC | [1] |
| | pan-Adenovirus | hexon | GCCACGGTGGGGTTTCTAAACTT | GCCCCAGTGGTCTTACATGCACATC | TGCACCAGACCCGGGCTCAG | [1] |
| | Rotavirus | NSP3 | ACCATCTWCACRTRACCCTCTATGAG | GGTCACATAACGCCCCTATAGC | AGTTAAAAGCTAACACTGTCAAA | [1] |
| Bacterium | *Campylobacter jejuni* | cadF | CTGCTAAACCATAGAAATAAAATTTCTCAC | CTTTGAAGGTAATTTAGATATGGATAATCG | CATTTTGACGATTTTTGGCTTGA | [1] |
| | *Clostridium dificile* | tcdB | GGTATTACCTAATGCTCCAAATAG | TTTGTGCCATCATTTTCTAAGC | CCTGGTGTCCATCCTGTTTC | [1] |
| | EAEC | aaiC | ATTGTCCTCAGGCATTTCAC | ACGACACCCCTGATAAACAA | TAGTGCATACTCATCATTTAAG | [1] |
| | | aatA | CTGGCGAAAGACTGTATCAT | TTTTGCTTCATAAGCCGATAGA | TGGTTCTCATCTATTACAGACAGC | [1] |
| | STEC1 | *stx1* | ACTTCTCGACTGCAAAGACGTATG | ACAAATTATCCCCTGWGCCACTATC | CTCTGCAATAGGTACTCCA | [1] |
| | STEC2 | *stx2* | CCACATCGGTGTCTGTTATTAACC | GGTCAAAACGCGCCTGATAG | TTGCTGTGGATATACGAGG | [1] |
| | EPEC | eae | CATTGATCAGGATTTTTCTGGTGATA | CTCATGCGGAAATAGCCGTTA | ATACTGGCGAGACTATTTCAA | [1] |
| | | bfpa | TGGTGCTTGCGCTTGCT | CGTTGCGCTCATTACTTCTG | CAGTCTGCGTCTGATTCCAA | [1] |
| | ETEC | LT | TTCCCACCGGATCACCAA | CAACCTTGTGGTGCATGATGA | CTTGGAGAGAAGAACCCT | [1] |
| | | STh | GCTAAACCAGYAGRGTCTTCAAAA | CCCGGTACARGCAGGATTACAACA | TGGTCCTGAAAGCATGAA | [1] |
| | | STp | TGAATCACTTGACTCTTCAAAA | GGCAGGATTACAACAAAGTT | TGAACAACACATTTTACTGCT | [1] |

| | | | | | | |
|---|---|---|---|---|---|---|
| | *Shigella*/EIEC | *ipaH* | CCTTTTCCGCGTTCCTTGA | CGGAATCCGGAGGTATTGC | CGCCTTTCCGATACCGTCTCTGCA | [1] |
| | *Salmonella enterica* | *invA* | TCGGGCAATTCGTTATTGG | GATAAACTGGACCACGGTGACA | AAGACAACAAAACCCACCGC | [2] |
| | *Vibrio cholerae* | *hlyA* | ATCGTCAGTTTGGAGCCAGT | TCGATGCGTTAAACACGAAG | ACCGATGCGATTGCCCAA | [2] |
| Protozoa | *Cryptosporidium hominis* | *LIB13* | TCCTTGAAATGAATATTTGTGACTCG | AAATGTGGTAGTTGCGGTTGAAA | CTTACTTCGTGGCGGCGT | [2] |
| | *Cryptosporidium parvum* | *LIB13* | TCCTTGAAATGAATATTTGTGACTCG | TTAATGTGGTAGTTGCGGTTGAAC | TATCTCTTCGTAGCGGCGTA | [2] |
| | *Giardia* spp. | 18S rRNA | GACGGCTCAGGACAACGGTT | TTGCCAGCGGTGTCCG | CCCGCGGCGGTCCCTGCTAG | [1] |
| | *Entamoeba histolytica* | 18S rRNA | ATTGTCGTGGCATCCTAACTCA | GCGGACGGCTCATTATAACA | TCATTGAATGAATTGGCCATTT | [1] |
| Helminth | *Ascaris lumbricoides* | *ITS1* | GCCACATAGTAAATTGCACACAAAT | GCCTTTCTAACAAGCCCAACAT | TTGGCGGACAATTGCATGCGAT | [2] |
| | *Trichuris trichiura* | 18S rRNA | TTGAAACGACTTGCTCATCAACTT | CTGATTCTCCGTTAACCGTTGTC | CGATGGTACGCTACGTGCTTACCATGG | [1] |
| | *Necator americanus* | *ITS2* | CTGTTTGTCGAACGGTACTTGC | ATAACAGCGTGCACATGTTGC | CTGTACTACGCATTGTATAC | [2] |
| | *Ancylostoma duodenale* | *ITS2* | GAATGACAGCAAACTCGTTGTTG | ATACTAGCCACTGCCGAAACGT | ATCGTTTACCGACTTTAG | [2] |
| Control | MS2 | *MS2g1* | TGGCACTACCCCTCTCCGTATTCAC | GTACGGGCGACCCCACGATGAC | CACATCGATAGATCAAGGTGCCTACAAGC | [1] |
| | bacterial 16S | | TGCAAGTCGAACGAAGCACTTTA | GCAGGTTACCCACGCGTTAC | CGCCACTCAGTCACAAA | [2] |


[1] Liu, J. et al. A Laboratory-Developed TaqMan Array Card for Simultaneous Detection of 19 Enteropathogens. J. Clin. Microbiol. 51, 472 LP – 480 (2013).
[2] Liu, J. et al. Optimization of Quantitative PCR Methods for Enteropathogen Detection. PLoS One 11, e0158199 (2016).
[3] Narayanan, J. et al. Quantitative Real-Time PCR Assays for Detection of Human Adenoviruses and Identification of Serotypes 40 and 41. Appl. Environ. Microbiol. 71, 3131–3136 (2005).

**S8: qPCR assay validation**

We tested preserved stools using a custom-developed TaqMan Array Card (TAC; ThermoFisher Scientific, Waltham, VA) – a compartmentalised probe-based quantitative polymerase chain reaction (qPCR) assay for enteric pathogen genes using individual assays validated in previously published literature in TAC format[9,10]. qPCR cycling conditions were also adapted from previous work[9,10] We further validated targets using synthetic nucleic acids (GeneArt, ThermoFisher Scientific) as positive controls (PCs). PC material for each individual assay was combined to a concentration of $10^{10}$ gene copies (GC)/uL. Two serial dilutions were run on the custom TAC: a high-concentration 10-fold dilution series ($10^{9}$ GC/uL to $10^{2}$ GC/uL) was used to determine range of the limit-of-quantification (LOQ) to order of magnitude; subsequently, a low-concentration 2-fold dilution series diluted within the determined LOQ range was used to estimate the delta-Rn threshold for each assay's LOQ.

**S9: Process evaluation indicators**

We documented process evaluation (PE) indicators to assess fidelty and adherence of intervention activities 28 months after the end of intervention roll-out. Fidelity was measured based on self-reported receipt of intervention activity, which included eight key nutrition activities: community dialogues (quarterly); caregiver group education course (monthly); village fairs (bi-annually); growth monitoring program (monthly); home health visits from VHSG (monthly); CCT with rolling enrolment (disbursed payments as participants met the various conditions); food vouchers (delivered once to CCT participants); and water filter vouchers (delivered once to CCT participants). Adherence was measured based on self-reported participation of intervention activities, which included household WASH practices and child nutrition behaviours.

**S10: Intervention adherence**

We assessed four key caregiver behaviours related to environmental hygiene: drinking and use of clean water, handwashing with soap and water at critical times, proper disposal of children's stools, and provision of safe play environments for children. Implementation programming encouraged safe handwashing behaviours as part of the "First 1,000 Days" activities and the nutrition CCT. Those in the nutrition-only arm (7%) and combined-intervention arm (9%) had greater awareness of critical handwashing times compared to those in the sanitation-only arm (4%) and control arm (4%), though levels were still low. There was a slightly higher prevalence of soap and water observed at handwashing stations in the combined-intervention (72%) and control (76%) arms than the nutrition-only (69%) and sanitation-only (70%) arms. We defined proper disposal of children's stools as discarding into a toilet/latrine or burying and considered discarding faeces into a drain, garbage or other solid waste, or leaving in the open to be improper disposal practices. Nutrition-only and combined intervention arms reported higher levels of proper disposal (71% and 74%, respectively) compared to the sanitation-only and control arms (65% and 68%, respectively). Few households were found to have safe play environments, defined as being free of observed human faeces, animal faeces, garbage/household waste, and sharp objects/other harms. 25% of households in the combined intervention arm had child play environments free of faeces observed by enumerators at the time of the household visit, compared to 21% in the nutrition-only, sanitation-only, and control arms. More households in the nutrition-only (78%) and combined-intervention (89%) arms brought children to health centres for monthly GMP visits than sanitation-only (23%) and control (33%) arms. There were no differences in breastfeeding behaviours between intervention and control arms, with 60-70% of each arm reporting continuous breastfeeding for children for the first two years. There were no statistically meaningful differences in dietary diversity score, minimum dietary diversity, and minimum meal frequency across the four arms.

#### S11: Adjusted analyses

Covariates were considered as potential confounders using a "common cause" approach[14] and based on the conceptual framework of literature-supported variables associated with diet and WASH conditions or nutritional status[15]. We also considered covariates that were found to be both associated with primary outcome measures and imbalanced across treatment arms before intervention delivery, of which only pre-intervention village-level sanitation coverage met the inclusion criteria. We calculated household wealth using an asset-based wealth index using methodology provided by the Demographic and Health Survey (DHS)[16], constructed using principal component analysis excluding WASH assets[17]. In the adjusted analyses, we included the following covariates, identified *a priori*: child sex (dichotomous), child age (continuous, in months), maternal age (continuous, in years), maternal education (ordinal, based on mother's highest level of education attended), number of household members (continuous), household wealth index quintile (ordinal), and community-level open defecation (OD) measured at prior to intervention rollout (continuous).

##### Effects of interventions on height/length and weight (Primary outcome (LAZ) and secondary outcomes (WAZ, WHZ)), comparing intervention arms to control and single intervention arms to combined intervention

| | | | | Compared to control arm | | Compared to combined intervention arm | |
|---|---|---|---|---|---|---|---|
| | N | Mean | SD | Unadjusted mean difference (95% CI) | Adjusted mean difference (95% CI) | Unadjusted mean difference (95% CI) | Adjusted mean difference (95% CI) |
| LAZ | | | | | | | |
| Nutrition-only | 798 | -0.95 | 1.16 | 0.08 (-0.01, 0.18) | 0.09 (-0.01, 0.19) | -0.02 (-0.12, 0.09) | 0.01 (-0.09, 0.11) |
| Sanitation-only | 777 | -1.09 | 1.23 | -0.05 (-0.16, 0.05) | -0.05 (-0.15, 0.05) | -0.16 (-0.27, -0.04) | -0.13 (-0.23, -0.02) |
| Combined | 1037 | -0.94 | 1.16 | 0.10 (0.01, 0.20) | 0.08 (-0.01, 0.17) | -- | -- |
| Control | 1443 | -1.04 | 1.20 | -- | -- | -- | -- |
| WAZ | | | | | | | |
| Nutrition-only | 815 | -0.95 | 1.29 | 0.10 (0.00, 0.19) | 0.10 (0.01, 0.20) | -0.02 (-0.12, 0.08) | 0.01 (-0.08, 0.11) |
| Sanitation-only | 792 | -1.04 | 1.13 | 0.01 (-0.07, 0.09) | 0.01 (-0.07, 0.09) | -0.10 (-0.20, -0.01) | -0.08 (-0.17, 0.00) |
| Combined | 1044 | -0.94 | 1.11 | 0.11 (0.03, 0.20) | 0.09 (0.01, 0.17) | -- | -- |
| Control | 1452 | -1.05 | 1.10 | -- | -- | -- | -- |
| WHZ | | | | | | | |
| Nutrition-only | 814 | -0.60 | 1.04 | 0.06 (-0.03, 0.15) | 0.06 (-0.02, 0.15) | -0.02 (-0.12, 0.08) | 0.00 (-0.09, 0.09) |
| Sanitation-only | 790 | -0.59 | 0.98 | 0.06 (-0.02, 0.14) | 0.05 (-0.03, 0.13) | -0.02 (-0.11, 0.07) | -0.01 (-0.10, 0.08) |
| Combined | 1043 | -0.58 | 1.03 | 0.08 (0.00, 0.16) | 0.06 (-0.02, 0.14) | -- | -- |
| Control | 1452 | -0.65 | 0.98 | -- | -- | -- | -- |
| Covariates in adjusted analyses include: child sex, child age, maternal age, maternal education, number of household members, household wealth index quintile, and community-level OD measured prior to intervention delivery | | | | | | | |

**Effects of intervention on child health outcomes, comparing intervention arms to control and single intervention arms to combined intervention.**

| | | | | Compared to control arm | | Compared to combined-intervention arm | |
|---|---|---|---|---|---|---|---|
| | N | Mean | SD | PR (95% CI) | aPR (95% CI) | PR (95% CI) | aPR (95% CI) |
| Stunted | | | | | | | |
| Nutrition-only | 801 | 0.15 | 0.36 | 0.84 (0.69, 1.03) | 0.83 (0.68, 1.02) | 0.93 (0.74, 1.15) | 0.90 (0.72, 1.12) |
| Sanitation-only | 782 | 0.21 | 0.40 | 1.12 (0.94, 1.33) | 1.11 (0.94, 1.31) | 1.23 (1.02, 1.49) | 1.20 (0.99, 1.46) |
| Combined | 1046 | 0.17 | 0.37 | 0.91 (0.76, 1.09) | 0.92 (0.77, 1.10) | -- | -- |
| Control | 1449 | 0.18 | 0.39 | -- | -- | -- | -- |
| Wasted | | | | | | | |
| Nutrition-only | 815 | 0.07 | 0.26 | 0.87 (0.65, 1.17) | 0.85 (0.63, 1.14) | 1.12 (0.80, 1.57) | 1.09 (0.78, 1.52) |
| Sanitation-only | 790 | 0.07 | 0.26 | 0.84 (0.62, 1.14) | 0.83 (0.61, 1.13) | 1.08 (0.76, 1.53) | 1.06 (0.75, 1.51) |
| Combined | 1052 | 0.07 | 0.25 | 0.78 (0.58, 1.04) | 0.78 (0.58, 1.05) | -- | -- |
| Control | 1457 | 0.08 | 0.28 | -- | -- | -- | -- |
| Underweight | | | | | | | |
| Nutrition-only | 816 | 0.15 | 0.35 | 0.85 (0.71, 1.03) | 0.84 (0.69, 1.02) | 1.04 (0.84, 1.29) | 0.99 (0.80, 1.24) |
| Sanitation-only | 792 | 0.17 | 0.38 | 1.00 (0.85, 1.19) | 1.00 (0.85, 1.17) | 1.22 (1.00, 1.49) | 1.18 (0.97, 1.44) |
| Combined | 1053 | 0.14 | 0.35 | 0.82 (0.68, 0.99) | 0.84 (0.70, 1.01) | -- | -- |
| Control | 1457 | 0.17 | 0.38 | -- | -- | -- | -- |
| Diarrhoea (7-day recall) | | | | | | | |
| Nutrition-only | 788 | 0.19 | 0.39 | 0.89 (0.74, 1.06) | 0.90 (0.76, 1.08) | 0.95 (0.78, 1.14) | 0.96 (0.80, 1.16) |
| Sanitation-only | 752 | 0.21 | 0.41 | 0.99 (0.84, 1.17) | 0.98 (0.83, 1.16) | 1.05 (0.88, 1.25) | 1.05 (0.87, 1.25) |
| Combined | 1018 | 0.20 | 0.40 | 0.94 (0.80, 1.11) | 0.94 (0.79, 1.11) | -- | -- |
| Control | 1411 | 0.21 | 0.41 | -- | -- | -- | -- |
| All-cause mortality | | | | | | | |
| Nutrition-only | 1574 | 0.03 | 0.16 | 1.55 (0.71, 3.39) | -- | 1.61 (0.68, 3.82) | -- |
| Sanitation-only | 1636 | 0.03 | 0.16 | 1.09 (0.50, 2.40) | -- | 1.13 (0.48, 2.68) | -- |
| Combined | 1932 | 0.03 | 0.16 | 0.96 (0.44, 2.10) | -- | -- | -- |
| Control | 2688 | 0.03 | 0.16 | -- | -- | -- | -- |
| Covariates in adjusted analyses include: child sex, child age, maternal age, maternal education, number of household members, household wealth index quintile, and community-level OD measured prior to intervention delivery | | | | | | | |

**Adjusted mean difference of detected bacteria, viruses, protozoa, and STHs, comparing intervention arms to control. Adjusted analyses controlled for the following covariates: child age, child sex, maternal age, maternal education, number of household members, household wealth quintile.**

| | | Compared to control arm | | | Compared to combined arm | |
|---|---|---|---|---|---|---|
| | N | Nutrition-only | Sanitation-only | Combined | Nutrition-only | Sanitation-only |
| Bacteria | 1406 | -0.04 | -0.09 | 0.02 | -0.07 | -0.11 |

| | | | | | | |
|---|---|---|---|---|---|---|
| | | (-0.22, 0.13) | (-0.27, 0.09) | (-0.15, 0.19) | (-0.25, 0.12) | (-0.30, 0.08) |
| Viruses | 786 | 0.08<br>(-0.01, 0.17) | -0.01<br>(-0.09, 0.07) | 0.06<br>(-0.02, 0.15) | 0.02<br>(-0.08, 0.12) | -0.07<br>(-0.16, 0.01) |
| Protozoa | 327 | 0.01<br>(-0.03, 0.05) | 0.01<br>(-0.03, 0.04) | 0.05<br>(-0.01, 0.11) | -0.04<br>(-0.10, 0.03) | -0.04<br>(-0.10, 0.02) |
| STH | 37 | 0.01<br>(-0.31, 0.32) | -0.10<br>(-0.42, 0.22) | 0.13<br>(-0.25, 0.52) | -0.13<br>(-0.43, 0.18) | -0.23<br>(-0.62, 0.16) |

**Adjusted prevalence ratios (aPR) of detected bacteria, viruses, protozoa, and STHs, comparing intervention arms to control. Adjusted analyses controlled for the following covariates: child age, child sex, maternal age, maternal education, number of household members, household wealth quintile**

| | Compared to control arm | | | Compared to combined arm | |
|---|---|---|---|---|---|
| | Nutrition-only | Sanitation-only | Combined | Nutrition-only | Sanitation-only |
| **Any bacterium** | 1.06 (1.01, 1.11) | 0.99 (0.93, 1.04) | 1.05 (1.00, 1.10) | 1.01 (0.96, 1.06) | 0.94 (0.89, 1.00) |
| *Camploybacter* spp. | 1.10 (0.90, 1.35) | 1.10 (0.91, 1.34) | 1.20 (1.01, 1.43) | 0.92 (0.75, 1.12) | 0.92 (0.76, 1.11) |
| *C.diff* | 1.43 (0.92, 2.22) | 0.98 (0.60, 1.59) | 1.21 (0.80, 1.83) | 1.18 (0.76, 1.83) | 0.81 (0.51, 1.29) |
| EAEC | 1.09 (0.98, 1.21) | 1.01 (0.90, 1.12) | 1.05 (0.96, 1.16) | 1.03 (0.93, 1.15) | 0.96 (0.86, 1.07) |
| EPEC | 0.95 (0.84, 1.07) | 0.87 (0.77, 0.99) | 0.94 (0.84, 1.04) | 1.01 (0.89, 1.15) | 0.93 (0.81, 1.07) |
| aEPEC | 0.96 (0.83, 1.12) | 0.87 (0.75, 1.02) | 0.87 (0.75, 1.00) | 1.11 (0.94, 1.31) | 1.01 (0.85, 1.20) |
| tEPEC | 0.61 (0.34, 1.08) | 0.59 (0.34, 1.00) | 0.94 (0.60, 1.47) | 0.65 (0.36, 1.18) | 0.62 (0.36, 1.10) |
| ETEC | 1.06 (0.84, 1.33) | 0.89 (0.70, 1.12) | 1.01 (0.82, 1.25) | 1.05 (0.83, 1.33) | 0.88 (0.69, 1.12) |
| ETEC-LT | 1.18 (0.91, 1.52) | 0.97 (0.75, 1.26) | 0.97 (0.76, 1.25) | 1.21 (0.92, 1.59) | 1.00 (0.75, 1.32) |
| ETEC-ST | 1.20 (0.82, 1.76) | 1.06 (0.71, 1.57) | 1.42 (1.01, 2.00) | 0.84 (0.59, 1.21) | 0.74 (0.51, 1.09) |
| ETEC-LT/ST | 1.89 (1.13, 3.16) | 1.62 (0.95, 2.77) | 1.74 (1.07, 2.84) | 1.08 (0.68, 1.73) | 0.93 (0.57, 1.52) |
| *Salmonella* spp. | 1.08 (0.69, 1.70) | 0.67 (0.40, 1.13) | 1.00 (0.65, 1.53) | 1.09 (0.68, 1.73) | 0.68 (0.40, 1.16) |
| EIEC/*Shigella* spp. | 0.56 (0.36, 0.86) | 0.86 (0.60, 1.22) | 0.95 (0.68, 1.32) | 0.59 (0.37, 0.92) | 0.90 (0.61, 1.32) |
| STEC | 1.22 (0.76, 1.94) | 0.92 (0.55, 1.52) | 1.46 (0.96, 2.22) | 0.83 (0.53, 1.31) | 0.63 (0.38, 1.03) |
| **Any virus** | 1.08 (0.93, 1.24) | 1.03 (0.89, 1.20) | 1.16 (1.02, 1.32) | 0.93 (0.80, 1.07) | 0.89 (0.77, 1.03) |
| Adenovirus | 1.80 (1.35, 2.40) | 1.30 (0.95, 1.78) | 1.36 (1.02, 1.83) | 1.32 (1.00, 1.74) | 0.95 (0.70, 1.30) |
| Enterovirus | 0.93 (0.76, 1.13) | 0.94 (0.77, 1.14) | 1.12 (0.94, 1.32) | 0.83 (0.68, 1.02) | 0.84 (0.69, 1.02) |
| **Any protozoa** | 0.90 (0.68, 1.20) | 1.01 (0.78, 1.30) | 1.14 (0.90, 1.45) | 0.79 (0.59, 1.06) | 0.88 (0.67, 1.16) |
| *Giardia* | 0.91 (0.68, 1.23) | 1.07 (0.82, 1.39) | 1.13 (0.88, 1.46) | 0.81 (0.59, 1.09) | 0.95 (0.72, 1.25) |
| Gene targets with <5% prevalence were omitted from PR analyses: *V.cholera*, astrovirus, norovirus, rotavirus, sapovirus, *Cryptosporidium*, *Entamoeba,* and all STHs (*Ascaris, Trichuris, Ancylostoma,* and *Necator*). | | | | | |

<u>Limitations</u>

This trial was limited in its capacity to measure intervention impacts due to our use of a single cross-sectional survey to retrospectively assess interventions delivered over the previous 28 months. This study included children born from 28 months before up to one month before the final measurement, with the primary outcome variable of age-adjusted linear growth on a continuous scale. As a result, children were exposed to varying levels of “maturity” of the interventions to which they are exposed.

**Impact of interventions on adjusted prevalence ratio of individual pathogens. Point estimates and 95% confidence intervals were determined using generalised log-linear Poisson models adjusting for covariates associated with each pathogen outcome: child age, child sex, maternal age, maternal education, number of household members, household wealth quintile.**

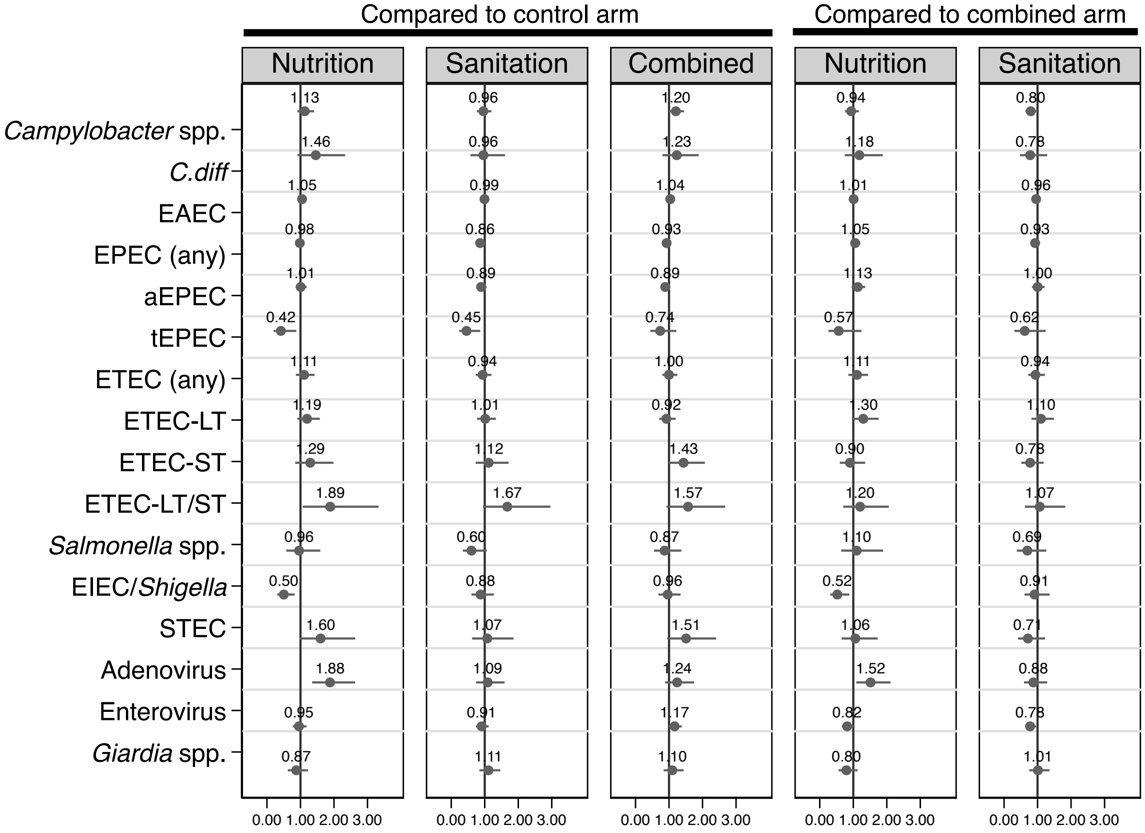

#### S12: Age-stratified pathogen prevalence

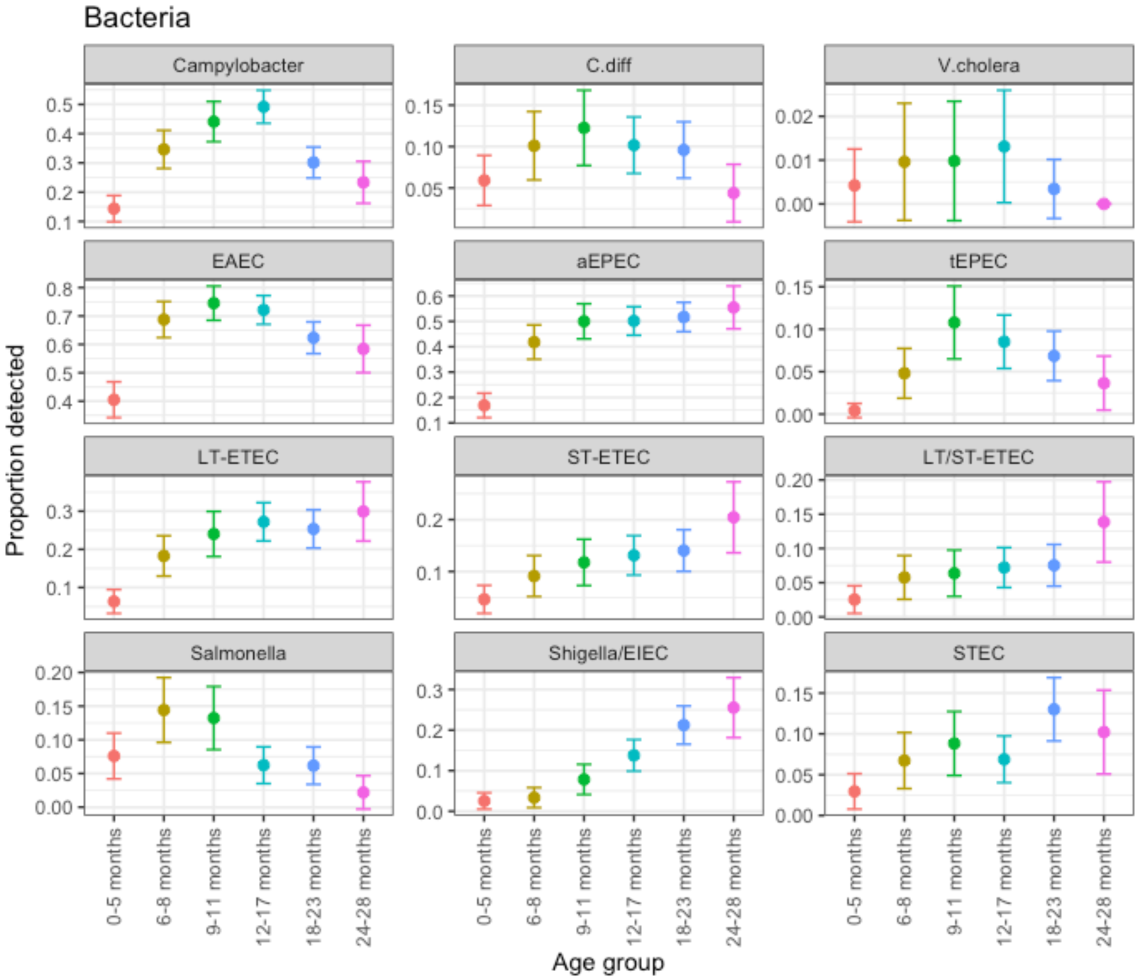

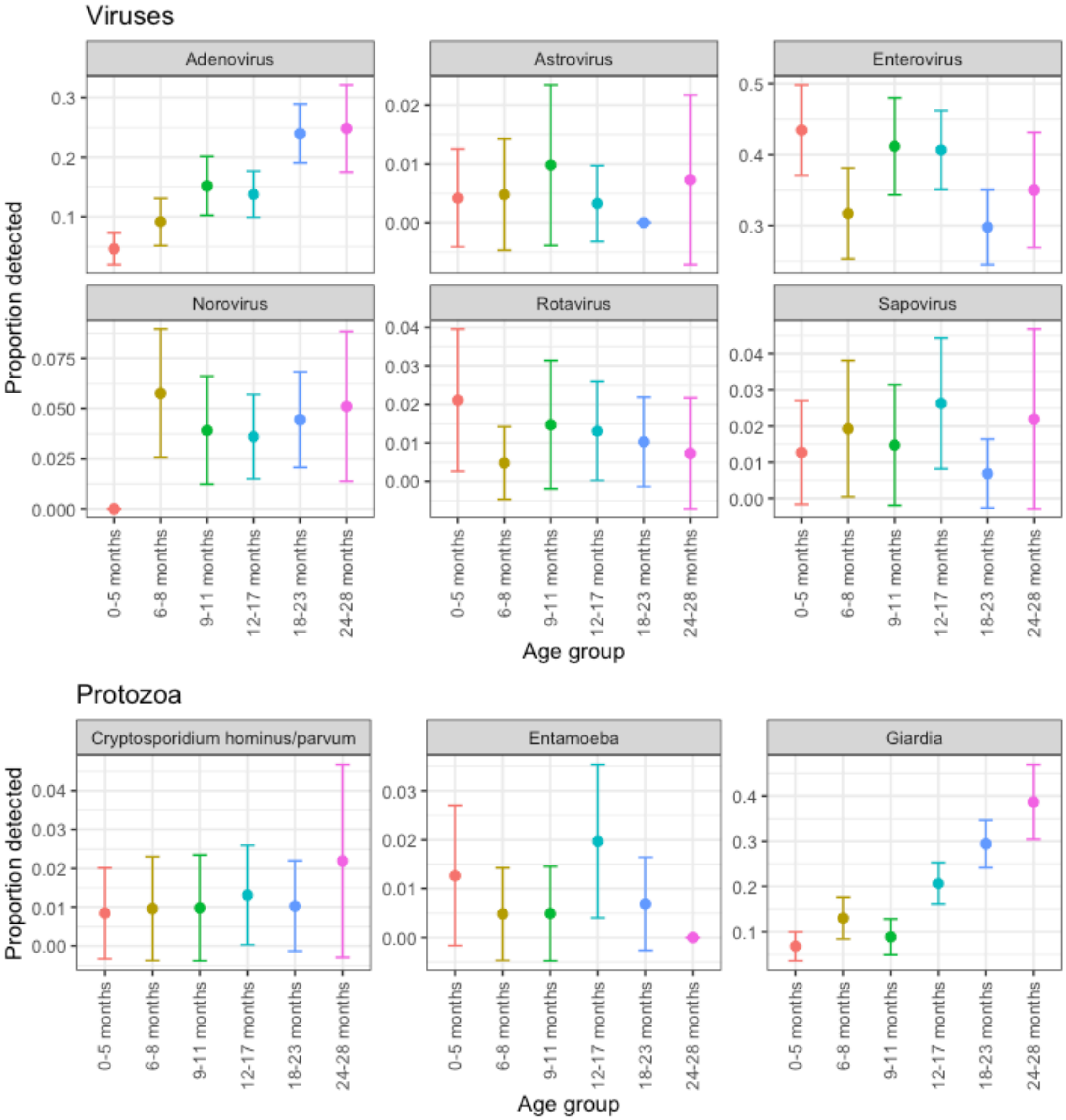
Viruses
Adenovirus
Astrovirus
Enterovirus
Norovirus
Rotavirus
Sapovirus
Proportion detected
0-5 months
6-8 months
9-11 months
12-17 months
18-23 months
24-28 months
Age group
Protozoa
Cryptosporidium hominus/parvum
Entamoeba
Giardia
Proportion detected
Age group

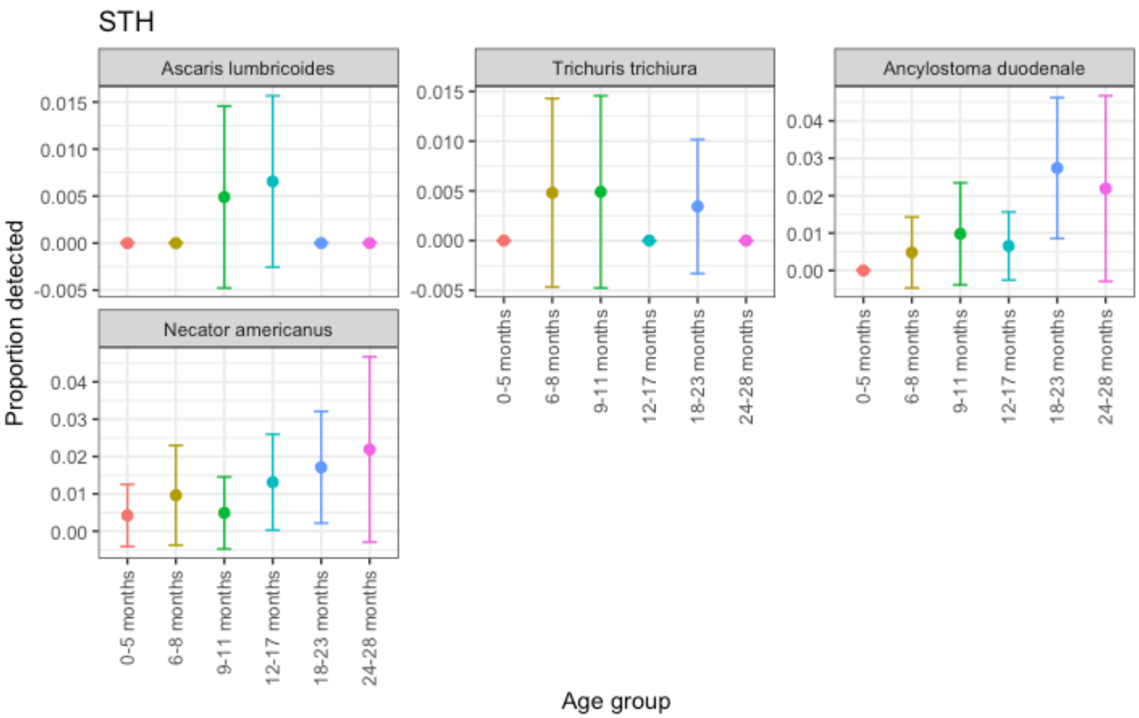
STH
Ascaris lumbricoides
Trichuris trichiura
Ancylostoma duodenale
Necator americanus
Proportion detected
Age group
0-5 months
6-8 months
9-11 months
12-17 months
18-23 months
24-28 months

##### S13: Pre-intervention sanitation coverage

| Treatment | Commune | % of households with improved drinking water source | % of households with access to improved sanitation facilities | % of households reporting open defecation practices | % of households with shared sanitation facilities | % of households reporting safe disposal of child stools |
|---|---|---|---|---|---|---|
| Nutrition | Chhnal Moan | 52% | 38% | 24% | 89% | 46% |
| Nutrition | Preah Phos | 67% | 41% | 48% | 85% | 58% |
| Nutrition | Prey Tralach | 86% | 29% | 43% | 55% | 53% |
| Nutrition | Robas Mongkol | 80% | 69% | 23% | 92% | 67% |
| Nutrition | Samraong | 67% | 48% | 33% | 84% | 48% |
| Nutrition | Sangvaeuy | 83% | 33% | 42% | 73% | 50% |
| Nutrition | Sdau | 75% | 54% | 25% | 88% | 63% |
| Nutrition | Ta Krei | 87% | 60% | 23% | 100% | 61% |
| Nutrition | Ta Pon | 80% | 80% | 7% | 92% | 75% |
| Nutrition | Ta Pung | 86% | 76% | 0% | 100% | 89% |
| Nutrition | Thipakdei | 94% | 64% | 21% | 91% | 65% |
| Sanitation | Basak | 60% | 20% | 73% | 75% | 18% |
| Sanitation | Chrouy Neang Nguon | 90% | 62% | 19% | 87% | 71% |
| Sanitation | Khnar Sanday | 95% | 43% | 50% | 100% | 41% |
| Sanitation | Lvea Krang | 89% | 33% | 44% | 75% | 40% |
| Sanitation | Pou Treay | 80% | 0% | 80% | 100% | 0% |
| Sanitation | Preaek Chik | 85% | 50% | 35% | 91% | 47% |
| Sanitation | Ruessei Krang | 48% | 27% | 58% | 90% | 25% |
| Sanitation | Srae Nouy | 72% | 36% | 53% | 87% | 46% |
| Sanitation | Ta Meun | 97% | 73% | 0% | 79% | 84% |
| Sanitation | Ta Yaek | 89% | 37% | 44% | 77% | 80% |
| Sanitation | Traeng | 100% | 54% | 33% | 87% | 71% |
| Sanitation | Varin | 81% | 48% | 48% | 91% | 27% |
| Sanitation | Yeang | 93% | 27% | 20% | 40% | 67% |
| Combined | Hab | 75% | 75% | 15% | 100% | 71% |
| Combined | Kaev Poar | 83% | 54% | 17% | 76% | 93% |
| Combined | Kakaoh | 79% | 46% | 21% | 79% | 71% |
| Combined | Kampong Preang | 94% | 83% | 6% | 94% | 70% |
| Combined | Kanhchor | 44% | 31% | 53% | 92% | 56% |
| Combined | Khnat | 92% | 67% | 6% | 92% | 73% |
| Combined | Mukh Paen | 100% | 39% | 33% | 70% | 50% |
| Combined | Prey Chruk | 100% | 42% | 27% | 82% | 63% |
| Combined | Slaeng Spean | 90% | 40% | 44% | 85% | 47% |

| | | | | | | |
|---|---|---|---|---|---|---|
| Combined | Srae Sdok | 75% | 37% | 35% | 74% | 49% |
| Combined | Svay sa | 81% | 62% | 14% | 81% | 80% |
| Combined | Ta Lou | 90% | 48% | 27% | 81% | 53% |
| Control | Anlong Reab | 85% | 31% | 38% | 71% | 88% |
| Control | Ballangk | 100% | 25% | 75% | 100% | 27% |
| Control | Chan Sar | 84% | 44% | 18% | 64% | 68% |
| Control | Chob Ta Trav | 100% | 40% | 60% | 100% | 36% |
| Control | Doun Ba | 95% | 59% | 27% | 93% | 81% |
| Control | Kantuot | 67% | 17% | 83% | 100% | 0% |
| Control | Kdei Run | 100% | 43% | 33% | 82% | 46% |
| Control | Khnar Pou | 79% | 63% | 13% | 83% | 65% |
| Control | Lveaeng Ruessei | 90% | 44% | 31% | 82% | 52% |
| Control | Mukh Reah | 76% | 33% | 38% | 70% | 53% |
| Control | Ou Ta Paong | 80% | 48% | 26% | 79% | 61% |
| Control | Prey Khpos | 100% | 80% | 7% | 96% | 90% |
| Control | Reaksmei Sangha | 70% | 30% | 26% | 58% | 71% |
| Control | Roung Kou | 96% | 26% | 56% | 70% | 21% |
| Control | Ruessei Lok | 71% | 33% | 50% | 80% | 33% |
| Control | Run Ta Aek | 86% | 24% | 38% | 63% | 44% |
| Control | Snuol | 96% | 26% | 52% | 70% | 54% |
| Control | Spean Tnaot | 84% | 62% | 7% | 82% | 71% |
| Control | Ta Loas | 100% | 67% | 15% | 78% | 70% |

**S14: CONSORT checklist for RCTs**

### CONSORT 2010 checklist of information to include when reporting a randomised trial*

| Section/Topic | Item No | Checklist item | Reported on page No |
|---|---|---|---|
| **Title and abstract** | | | |
| | 1a | Identification as a randomised trial in the title | 1 |
| | 1b | Structured summary of trial design, methods, results, and conclusions (for specific guidance see CONSORT for abstracts) | 1, 2 |
| **Introduction** | | | |
| Background and objectives | 2a | Scientific background and explanation of rationale | 3 |
| | 2b | Specific objectives or hypotheses | 3,4 |
| **Methods** | | | |
| Trial design | 3a | Description of trial design (such as parallel, factorial) including allocation ratio | 4 |
| | 3b | Important changes to methods after trial commencement (such as eligibility criteria), with reasons | 4, Supplementary Material |
| Participants | 4a | Eligibility criteria for participants | 4, Supplementary Material |
| | 4b | Settings and locations where the data were collected | 4 |
| Interventions | 5 | The interventions for each group with sufficient details to allow replication, including how and when they were actually administered | 4, Supplementary Material |
| Outcomes | 6a | Completely defined pre-specified primary and secondary outcome measures, including how and when they were assessed | 5 |
| | 6b | Any changes to trial outcomes after the trial commenced, with reasons | n/a |
| Sample size | 7a | How sample size was determined | 5, Supplementary Material |
| | 7b | When applicable, explanation of any interim analyses and stopping guidelines | n/a |
| Randomisation: | | | |
| Sequence generation | 8a | Method used to generate the random allocation sequence | 4 |
| | 8b | Type of randomisation; details of any restriction (such as blocking and block size) | 4 |
| Allocation concealment mechanism | 9 | Mechanism used to implement the random allocation sequence (such as sequentially numbered containers), describing any steps taken to conceal the sequence until interventions were assigned | 4 |
| Implementation | 10 | Who generated the random allocation sequence, who enrolled participants, and who assigned participants to interventions | 4 |

| | | | |
|---|---|---|---|
| Blinding | 11a | If done, who was blinded after assignment to interventions (for example, participants, care providers, those assessing outcomes) and how | 4 |
| | 11b | If relevant, description of the similarity of interventions | n/a |
| Statistical methods | 12a | Statistical methods used to compare groups for primary and secondary outcomes | 5 |
| | 12b | Methods for additional analyses, such as subgroup analyses and adjusted analyses | 5, Supplementary Material |
| **Results** | | | |
| Participant flow (a diagram is strongly recommended) | 13a | For each group, the numbers of participants who were randomly assigned, received intended treatment, and were analysed for the primary outcome | 6 |
| | 13b | For each group, losses and exclusions after randomisation, together with reasons | 6 |
| Recruitment | 14a | Dates defining the periods of recruitment and follow-up | 4 |
| | 14b | Why the trial ended or was stopped | n/a |
| Baseline data | 15 | A table showing baseline demographic and clinical characteristics for each group | Table 2 |
| Numbers analysed | 16 | For each group, number of participants (denominator) included in each analysis and whether the analysis was by original assigned groups | 6 |
| Outcomes and estimation | 17a | For each primary and secondary outcome, results for each group, and the estimated effect size and its precision (such as 95% confidence interval) | 6-8, Figure 3, Figure 4, Tables 6-10 |
| | 17b | For binary outcomes, presentation of both absolute and relative effect sizes is recommended | Tables 6-10 |
| Ancillary analyses | 18 | Results of any other analyses performed, including subgroup analyses and adjusted analyses, distinguishing pre-specified from exploratory | Supplementary Material |
| Harms | 19 | All important harms or unintended effects in each group (for specific guidance see CONSORT for harms) | n/a |
| **Discussion** | | | |
| Limitations | 20 | Trial limitations, addressing sources of potential bias, imprecision, and, if relevant, multiplicity of analyses | 9-10 |
| Generalisability | 21 | Generalisability (external validity, applicability) of the trial findings | 10-11 |
| Interpretation | 22 | Interpretation consistent with results, balancing benefits and harms, and considering other relevant evidence | 9-11 |
| **Other information** | | | |
| Registration | 23 | Registration number and name of trial registry | 12 |
| Protocol | 24 | Where the full trial protocol can be accessed, if available | 12 |
| Funding | 25 | Sources of funding and other support (such as supply of drugs), role of funders | 12 |

*We strongly recommend reading this statement in conjunction with the CONSORT 2010 Explanation and Elaboration for important clarifications on all the items. If relevant, we also recommend reading CONSORT extensions for cluster randomised trials, non-inferiority and equivalence trials, non-pharmacological treatments, herbal interventions, and pragmatic trials. Additional extensions are forthcoming: for those and for up to date references relevant to this checklist, see www.consort-statement.org.
